# STDP-inspired temporal transition modeling for adaptive clinical risk prediction from electronic health records

**DOI:** 10.64898/2026.06.04.26354919

**Authors:** Liyuan Gong, Nitish Aswani, Peter Shahinnian, Jeong Yun Yang, Despina Kontos, Gulam Manji, Stella Kang, Chin Hur

## Abstract

Electronic health record (EHR) prediction models often summarize longitudinal histories as static patient-level features, which may omit potentially informative event ordering. We developed a simplified spike-timing-dependent plasticity (STDP)-inspired framework that represents asynchronous EHR data as sparse, directional transition features. The approach encodes whether one clinical event precedes another within prespecified temporal windows, preserving event identity, directionality, and approximate timing while retaining feature-level interpretability. We evaluated this framework in two retrospective prediction tasks with different temporal scales: incident acute kidney injury (AKI) prediction in 17,351 MIMIC-IV ICU stays and early postoperative recurrence prediction in 713 CUMC patients with pancreatic ductal adenocarcinoma (PDAC). Models were compared with static burden features (demographics, comorbidities, raw lab measurements) and in addition with STDP transitional feature sets using patient-level cross-validation and rolling prediction horizons. In AKI, a calibrated STDP ensemble model showed higher discrimination than static burden alone at the 24-hour decision snapshot for AKI by 72 hours, with AUROC 0.838 versus 0.800, and at 48 hours for near-term AKI prediction, with AUROC 0.868 versus 0.827. In PDAC, STDP transition features modestly improved Day –30 preoperative recurrence prediction, with AUROC 0.611 versus 0.587 and AUPRC 0.323 versus 0.318 for static burden and showed similar performance at Day 0 (7 days before recorded surgery date), with AUROC 0.681 and AUPRC 0.363. Decision-curve and feature analyses suggested that selected temporal transitions were clinically interpretable across renal, inflammatory, hepatobiliary, hematologic, glycemic, and nutritional trajectories. These findings suggest that STDP-inspired transition features may provide a practical, interpretable way to incorporate temporal ordering into EHR-based risk prediction across both acute and longitudinal settings.

## 1. Introduction

Clinical prediction models based on electronic health record data are increasingly used for point-of-care risk stratification, including identifying patients at risk for adverse outcomes such as mortality, readmission, prolonged length of stay, clinical deterioration, and adverse drug events.^1^ These models are also being embedded into clinical workflows to support real-time surveillance and earlier intervention for high-risk patients.^2^ Many rely on structured EHR data available at scale, including diagnosis codes, procedures, laboratory values, medication exposures, administrative/encounter data, and utilization-related variables.^3^ Although such approaches can support scalable and reproducible risk modeling, they may collapse clinically meaningful temporal structure. As a result, two patients with the same set of diagnoses and laboratory abnormalities may receive similar model representations even if one patient developed abnormalities in a rapidly worsening sequence and the other accumulated them over a long, clinically stable period.

Temporal EHR data are often preferred for prediction tasks because they can provide more complete representations of a patient’s pathophysiology than static data, but temporal modeling introduces challenges around time-window definition, irregularity, heterogeneity, sparsity, and reporting.^4,5^ In practice, clinicians repeatedly update their assessment as new symptoms, diagnoses, laboratory abnormalities, physiologic changes, procedures, and treatment responses appear. The order in which events occur can matter as much as their presence. In AKI, for example, Kidney Disease: Improving Global Outcomes (KDIGO) defines kidney injury using both serum creatinine rise and reduced urine output, reflecting that renal dysfunction may be detected through multiple evolving physiologic signals rather than a single laboratory value alone.^6^ In PDAC, clinically meaningful risk can also evolve over time. Serial CA19-9 dynamics have been associated with recurrence risk, and rising CA19-9 may precede radiographic recurrence in postoperative surveillance.^7,8^ Biliary obstruction and jaundice can emerge as pancreatic tumors progress and may affect both treatment timing and biomarker interpretation.^9^ These examples suggest that event order, such as liver or biliary abnormalities followed by jaundice-related care, or hematologic and inflammatory abnormalities followed by nutritional decline, may carry prognostic information beyond unordered cumulative event counts.

Temporal EHR models have attempted to address this limitation using time bins, recurrent neural networks, transformers, sequence embeddings, and large clinical foundation models. Prior work has treated EHR data as ordered longitudinal event sequences, including sequence-of-visits representations, recurrent models such as Doctor AI and DeepCare, transformer-based models such as BEHRT, and more recent pretrained or generative EHR models for forecasting future diagnoses and clinical events.^10–13^ These studies support the premise that the order and timing of clinical events contain predictive information beyond cumulative event burden. However, many temporal models require large datasets, substantial computational resources, and architectures that encode time into latent representations that can be difficult to interpret clinically. Reviews of temporal deep learning for EHR data have also highlighted persistent challenges related to irregular sampling, heterogeneity, sparsity, and model opacity.^14^ In addition, temporal models are often evaluated as endpoint predictors rather than as rolling decision tools, so strong performance near an outcome may reflect proximal clinical deterioration rather than earlier actionable warning.

Spike-timing-dependent plasticity (STDP), a temporally asymmetric form of Hebbian learning, offers a biologically inspired way to represent event order while preserving feature-level interpretability.^15^ In neurobiology, the relative timing of paired events influences whether their association is strengthened or weakened. Beyond neuroscience, STDP-inspired learning rules have been used in spiking neural networks and neuromorphic systems for temporal pattern recognition, event-based visual classification, and efficient local feature learning, where information is encoded through the timing of discrete events rather than through continuously sampled measurements.^16,17^ These applications motivate STDP as a natural computational analogy for irregular EHR data, in which diagnoses, laboratory abnormalities, treatments, and procedures occur asynchronously and may carry prognostic information through their temporal ordering.

In this study, we adapted the STDP timing principle to structured EHR data and evaluated it in two complementary settings: incident AKI prediction in MIMIC-IV, a dense intensive-care benchmark, and early postoperative recurrence prediction in PDAC, a sparse longitudinal oncology surveillance task. In PDAC, all patients had the same underlying cancer diagnosis, making recurrence prediction a within-disease prognostic task rather than cancer-versus-control classification. We compared static burden models, STDP transition-only models, and STDP-enhanced models using patient-level cross-validation and rolling prediction horizons. We further assessed discrimination, calibration, decision curves, high-specificity operating points, and feature attribution to evaluate incremental value and interpretability.

## 2. Materials and methods

### 2.1. Study design and data sources

We conducted a retrospective modeling study to evaluate an STDP-inspired temporal transition framework for adaptive clinical risk prediction from structured EHR data. Two prediction tasks were selected to test the framework across different temporal scales and observation densities: incident acute kidney injury (AKI) prediction in MIMIC-IV 3.1^18^ intensive care unit data and early postoperative recurrence prediction in an institutional PDAC cohort from Columbia University Irving Medical Center (CUMC). The AKI task represented a dense acute-care setting with frequent laboratory and physiologic measurements, whereas the PDAC task represented a sparse longitudinal oncology surveillance setting. The retrospective study protocol using de-identified patient data was reviewed and approved by the CUMC Institutional Review Board (IRB protocols: AAAS1319).

### 2.2. Cohort definitions and outcomes

For the AKI task, we identified adult ICU stays with sufficient 24 hours creatinine and follow-up (≥72 hours) data to ascertain incident AKI. AKI occurring before or at the prediction snapshot was excluded to ensure that models predicted future rather than already established AKI. AKI outcomes were defined using creatinine-based and/or urine-output-based criteria consistent with KDIGO principles,^19^ with baseline creatinine estimated from available pre-ICU or early-ICU measurements according to the cohort algorithm. The primary AKI prediction tasks used information available during the first 24 hours of ICU admission to predict AKI over subsequent 48-hour and 72-hour horizons, with controls patients who were selected as AKI free up to the same snapshot and did not develop AKI by 72h from ICU admission. For the PDAC task, we assembled an institutional cohort of patients with PDAC who had adequate preoperative EHR (≥ 1yr) history and postoperative follow-up (≥ 6 months) for recurrence ascertainment. The outcome was early postoperative recurrence within 6 months, confirmed by pathological findings. Patients with insufficient follow-up records for recurrence ascertainment were excluded. The index date was based on curative-intent surgery or the closest available surgical/index proxy. Because structured OMOP procedure codes may be recorded after the actual operative date, Day 0 was defined as 7 days before the recorded surgical procedure code to reduce potential leakage from perioperative or postoperative documentation. Day –30 was defined as a priori as the primary preoperative decision horizon.

### 2.3. Clinical event representation

Structured EHR records were mapped into timestamped clinical event families before feature construction. Standard OMOP diagnosis codes were grouped into clinically interpretable condition categories. Abnormal laboratory measurements were defined using clinically meaningful thresholds (Table S1). For ablation analysis, we grouped heterogeneous laboratory values to be represented as physiological domains such as renal, hepatobiliary, inflammatory, hematologic, glycemic, electrolyte, and nutritional axes. For AKI, event representations emphasized ICU measurements, renal function, urine output, hemodynamic context, laboratory density, and short-horizon physiologic change. For PDAC, event representations emphasized preoperative diagnosis groups, abnormal laboratory, hepatobiliary and gastrointestinal symptoms, inflammatory and nutritional trajectories, and utilization frequency. For each prediction snapshot, only events occurring before the horizon-specific cutoff were used. We did not include treatment or procedures in the predictive features to simply the framework.

### 2.4. STDP-inspired transition feature construction

For each patient-snapshot, events were sorted chronologically and converted into sparse directional transition features. Motivated by pair-based spike-timing-dependent plasticity models, in which the strength of association between two spikes depends on their relative timing,^20,21^ we adapted an exponential timing window to ordered EHR event pairs. For an ordered pair of event types (*i*) and (*j*), occurring at times (*t*_*i*_) and (*t*_*j*_), a transition contribution was assigned only when event i preceded event j within a prespecified lag window. Specifically, a pair was retained when the elapsed time between the two events was greater than zero and less than or equal to the maximum allowable lag L. For qualifying event pairs, the transition contribution was weighted using an exponential decay function, where *τ* controls how rapidly temporally distant event pairs are down-weighted. Directionality was preserved, so *i*→*j* and *j*→*i* were treated as distinct features. Contributions from repeated qualifying pairs were aggregated within each patient-snapshot to form transition scores. Lag and decay parameters were treated as empirical hyperparameters rather than fixed biological constants. Candidate lag and decay settings were compared during model development, and final frozen models retained transition families that improved validation performance while excluding broad or noisy transition families. The transition feature construction is summarized in Equations (1)-(3):

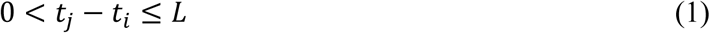

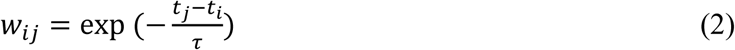

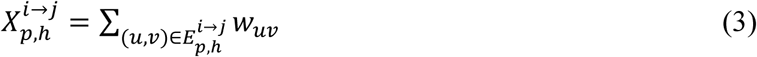

where 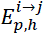 denotes the set of qualifying ordered event pairs of type *i*→*j* for patient p at prediction horizon h. And 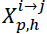 denotes the resulting aggregated transition score.

### 2.5. Baseline and STDP-enhanced model families

We compared several model families to isolate the incremental value of temporal transition features. Static burden baselines summarized cumulative pre-cutoff information, including condition counts, abnormal laboratory-axis counts, utilization summaries, demographic variables, comorbidity indicators, and recent or baseline laboratory measurements when applicable. Single-STDP transition models used directional transition features as the primary temporal representation. STDP-enhanced models concatenated static burden features with STDP transition features to evaluate whether event ordering added predictive information beyond cumulative burden alone. Final calibrated STDP ensembles combined selected stable component models with calibration procedures to improve discrimination and probability estimation. Across model families, prediction heads included logistic regression, elastic net, XGBoost, and LightGBM.

### 2.6. Rolling prediction design and leakage prevention

Rolling prediction snapshots were generated separately for each task. For PDAC, prediction horizons were Day –90, –60, –30, –14, –7, –3, and Day 0 relative to the surgery/index date, with Day –30 serving as the primary clinical decision horizon and Day 0 used as a sustained-performance horizon. The recurrence outcome label was fixed at the patient level, but each horizon used a distinct preoperative feature cutoff. Day 0 was defined as 7 days before the recorded surgical procedure code, as described above, to reduce potential leakage from perioperative or postoperative documentation. For AKI, rolling ICU snapshots were generated at clinically relevant time points, including 6, 12, 24, 36, 48, and 72 hours after ICU admission or relative to the target prediction horizon. For every snapshot, feature construction was restricted to information available before the cutoff, and patients or ICU stays with the outcome already present at or before the snapshot were excluded from future-event prediction. Cross-validation and test-set evaluation were performed at the patient or ICU-stay level, ensuring that all rolling snapshots from the same individual remained in the same fold and preventing leakage across horizons. Final frozen model configurations were then evaluated across rolling horizons using the same patient-level split structure.

### 2.7. Performance evaluation, robustness, and statistical analysis

Model performance was evaluated using discrimination, calibration, and clinical utility metrics. Primary discrimination metrics were AUROC and AUPRC. Probability accuracy was assessed using Brier score and calibration curves. Clinically relevant operating points were summarized using sensitivity and positive predictive value at high-specificity thresholds, with 90% specificity selected as the primary operating point. High-specificity performance was emphasized because PDAC recurrence prediction may inform preoperative surgical decision-making, where false-positive high-risk classification could alter surgical planning, trigger additional testing, or intensify treatment discussions. Decision-curve analysis was used to estimate net benefit across threshold probabilities rather than to select a single universal intervention threshold. For AKI, threshold probabilities were interpreted in the context of lower-burden interventions such as intensified monitoring, nephrotoxin review, hemodynamic reassessment, or repeat laboratory testing. For PDAC, moderate-to-high threshold regions were emphasized because recurrence-risk estimates could influence tumor-board discussion, imaging, treatment sequencing, or surveillance intensity. Confidence intervals were estimated using bootstrap resampling at the patient or ICU-stay level. Paired AUROC comparisons between STDP-enhanced models and static burden baselines were performed using paired DeLong tests when paired prediction vectors were available. Paired bootstrap deltas were used for AUPRC, Brier score, sensitivity, and positive predictive value. Robustness analyses evaluated whether STDP performance was explained by observation opportunity. PDAC models were assessed across minimum prior-record requirements, prior-record-length strata, and encounter/event/laboratory-density adjustment. AKI robustness was evaluated using first-24-hour laboratory measurement intensity. Interpretability analyses used model coefficients, XGBoost gain, transition-signal evolution, high-risk decile enrichment, and exploratory transition-velocity summaries.

## 3. Results

### 3.1. STDP transition modeling framework

We developed an STDP-inspired framework to represent longitudinal EHR data as sparse, directional clinical transition features (Fig. 1). Patient timelines were mapped into ordered clinical events, including diagnosis groups, abnormal laboratory axes, treatment exposures when available, and utilization markers. Laboratory measurements were converted into clinically meaningful abnormality axes, allowing heterogeneous values to be represented as interpretable physiologic domains such as renal, hepatobiliary, inflammatory, hematologic, glycemic, and nutritional trajectories. For each task and prediction horizon, feature construction was restricted to information available before the horizon-specific cutoff. In PDAC, rolling snapshots were generated at Day –90, –60, –30, –14, –7, –3, and 0 relative to surgery or index date, with Day –30 defined a priori as the primary surgical decision horizon. In AKI, snapshots were generated at clinically relevant ICU time points before AKI onset or matched control time.

**Fig 1.**
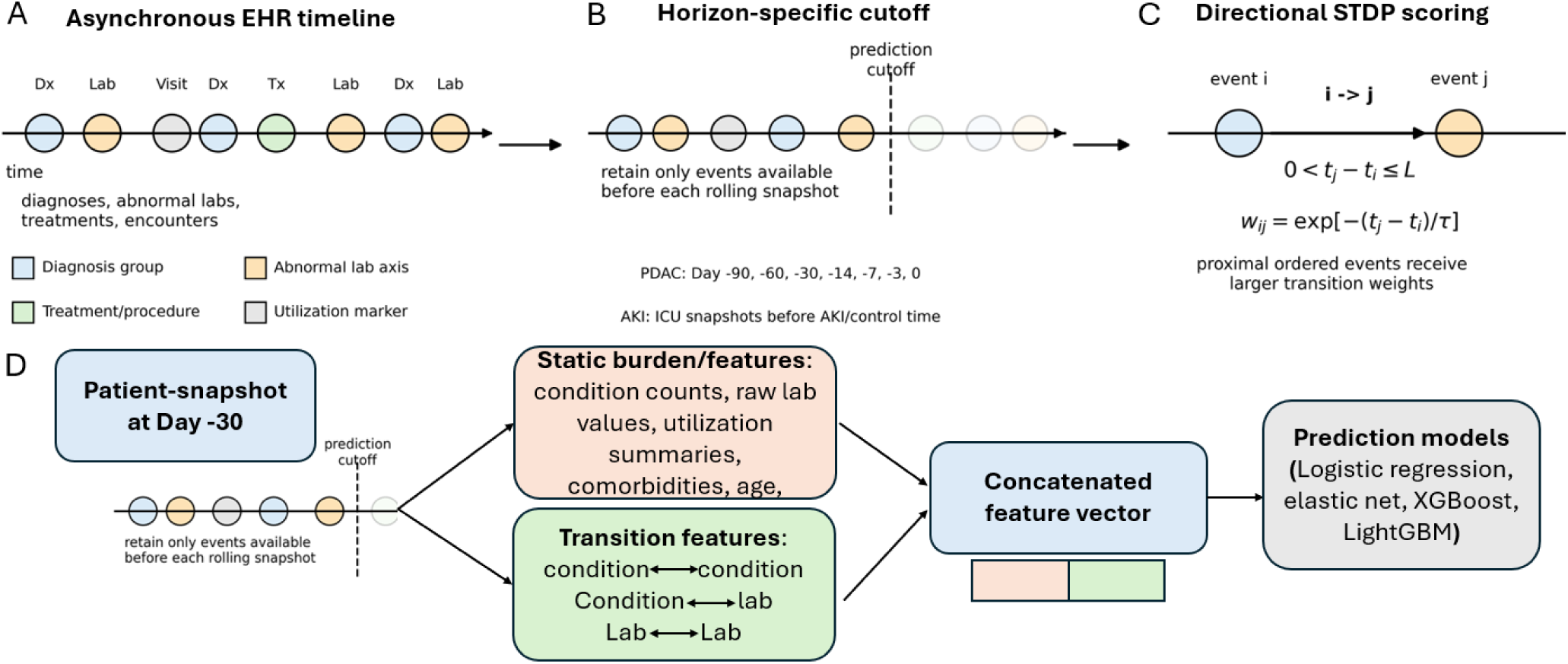
STDP-inspired EHR transition modeling framework. A, Irregular EHR histories are represented as asynchronous clinical timelines containing diagnosis groups, abnormal laboratory axes, treatment or procedure exposures, and utilization markers. B, for each rolling prediction snapshot, a horizon-specific cutoff is applied so that only events available before the prediction time are used for feature construction. PDAC recurrence models used preoperative snapshots from Day –90 to Day 0, with Day –30 defined as the primary surgical decision horizon; AKI models used ICU snapshots before AKI onset or matched control time. C, Directional STDP-inspired transition scoring encodes whether event (i) precedes event (j) within a prespecified lag window, with more proximally ordered transitions receiving larger weights. D, for each patient-snapshot, pre-cutoff information is transformed into feature blocks, including static burden features and directional transition features such as condition-to-condition, condition-to-laboratory, and laboratory-to-condition trajectories. These feature blocks are concatenated into a patient-snapshot feature vector and used by the prediction model to estimate rolling risk.

For each patient-snapshot, static burden features and directional transition features were constructed from the same pre-cutoff history. Static burden features summarized cumulative clinical information, including condition counts, laboratory abnormalities, utilization summaries, comorbidities, and demographic variables when available. STDP-inspired transition features encoded temporally ordered event pairs, including condition-to-condition, condition-to-laboratory, and laboratory-to-condition trajectories, with proximal ordered transitions contributing more strongly than distant co-occurrences. The resulting feature blocks were concatenated and evaluated using static burden, STDP transition-only, and STDP-enhanced models. All models were trained and evaluated using patient-level cross-validation, ensuring that rolling snapshots from the same patient remained in the same fold and preventing information leakage across prediction horizons.

### 3.2. Cohort characteristics

The AKI cohort included 17,351 ICU stays, of which 3,532 developed incident AKI, corresponding to an event rate of 20.4%. The median age was 67.0 years [IQR, 55.0-78.0], and 7,676 patients or stays were female (44.2%). The PDAC recurrence cohort included 713 patients with adequate postoperative follow-up, of whom 164 developed recurrences within 6 months, corresponding to an event rate of 23.0%. The median age was 73.1 years [IQR, 63.9-80.3], and 346 patients were female (48.5%). The two cohorts were selected to test the same temporal transition framework across different clinical time scales and data densities. The AKI task used early ICU information from the first 24 hours to predict AKI onset over the subsequent 48-72 hours, with rolling snapshots at 6, 12, 24, 36, 48, and 72 hours. The PDAC task used longitudinal preoperative EHR history to predict postoperative recurrence within 6 months, with rolling snapshots at Day –90, –60, –30, –14, –7, –3, and Day 0 relative to the surgery/index date. Day –30 was defined as the primary preoperative decision horizon for recurrence prediction.

### 3.3. AKI benchmark prediction performance

We first evaluated the STDP-inspired framework in MIMIC-IV AKI prediction. The primary analysis used information available during the first 24 hours of ICU admission to predict incident AKI over subsequent 48-hour and 72-hour horizons. In the held-out test set for AKI by 48 hours, the calibrated STDP ensemble achieved the highest discrimination, with AUROC 0.868 [95% CI, 0.851-0.884], compared with 0.827 [0.807-0.843] for the static burden baseline and 0.832 [0.815-0.850] for the single STDP transition model (Fig. 2A; Table 2). The ensemble also improved AUPRC from 0.486 [0.440-0.529] to 0.581 [0.537-0.625], reduced Brier score from 0.147 [0.141-0.153] to 0.123 [0.117-0.129], and increased sensitivity at 90% specificity from 0.507 [0.455-0.554] to 0.609 [0.559-0.656]. Decision-curve analysis showed higher net benefit for the calibrated STDP ensemble across most threshold probabilities, indicating that the improvement in discrimination translated into improved clinical utility (Fig. 2B).

**Figure 2.**
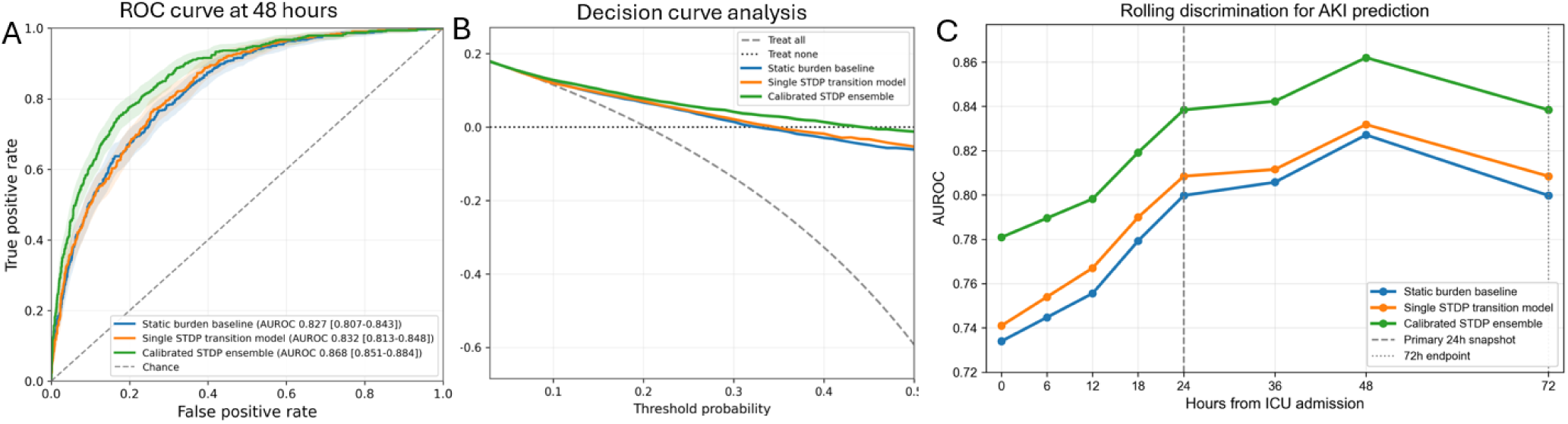
STDP-enhanced rolling prediction of incident AKI. A, ROC curves for AKI prediction at the 48-hour horizon. B, Decision-curve analysis comparing net benefit across threshold probabilities. Rolling AUROC across ICU snapshots, with the primary 24-hour snapshot and 72-hour horizon marked. The calibrated STDP ensemble outperformed the static burden baseline and single STDP transition model across discrimination and decision-curve analyses.

**Table 1.** Summary of cohort characteristics and prediction task definitions. **Footnote:** Values are reported as n (%), median [IQR], or cohort descriptors. For MIMIC-IV, the unit of analysis was ICU stay/patient snapshot. For PDAC, the unit of analysis was patient/preoperative prediction horizon. In the PDAC cohort, Day 0 was defined as 7 days before the recorded surgical procedure code because procedure codes may be entered after the actual operative date; this definition was used to reduce leakage from perioperative or postoperative documentation. Patients with insufficient postoperative follow-up for recurrence ascertainment, defined as follow-up records ≤6 months, were excluded from the recurrence cohort. Race categories were derived from structured source values.

| Characteristic | MIMIC-IV AKI cohort | PDAC recurrence cohort |
| --- | --- | --- |
| Data source | MIMIC-IV 3.1 ICU | CUMC Institutional PDAC cohort |
| Prediction task | Incident AKI prediction | Early recurrence prediction |
| Unit of analysis | ICU stay / patient snapshot | Patient / preoperative horizon |
| Number of patients | 17,351 | 713 |
| Number of ICU stays / index encounters | 17,351 | 713 |
| Positive cases | 3,532 | 164 |
| Event rate | 20.4% | 23.0% |
| Age, median or mean | 67.0 [55.0, 78.0] | 73.1 [63.9-80.3] |
| Female sex, n (%) | 7,676 (44.2%) | 346 (48.5%) |
| White | 10423 (60.1%) | 424 (59.5%) |
| Black/African American | 4233 (24.4%) | 119 (16.7%) |
| Asian ancestry | 508 (2.9%) | 28(3.9%) |
| Other or unknown | 2,187 (12.6%) | 142 (19.9%) |
| Observation window | ICU 0-24h for primary model | Preoperative EHR history |
| Primary prediction horizon | AKI onset 48-72h | Postoperative within 6 months |
| Rolling snapshots | 6, 12, 24, 36, 48, 72h | Day -90, -60, -30, -14, -7, -3, 0 |
| Primary decision horizon | ICU 24h → next 48h | Day -30 before surgery/index |
| Key exclusions | Early AKI $\leq 24h$ or ≤ snapshot; inadequate creatinine/follow-up | Follow up records $\leq 6$ months |

**Table 2.**
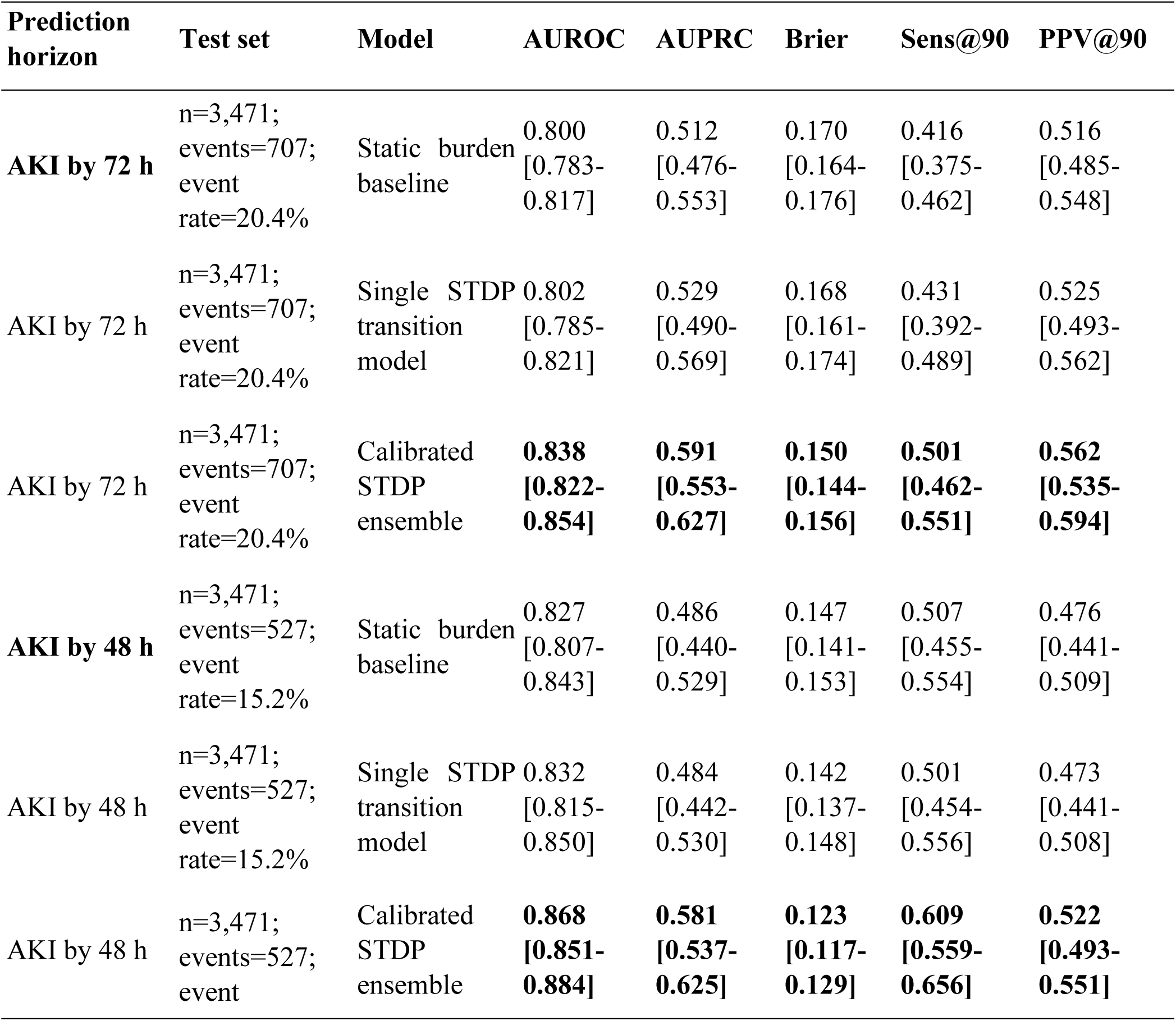
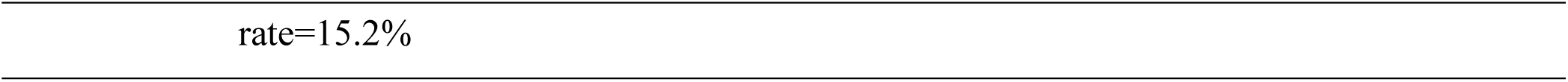
AKI prediction performance at 48-hour and 72-hour horizons.

Rolling evaluation across ICU snapshots showed that discrimination increased as additional ICU data accumulated, but the calibrated STDP ensemble remained consistently above both the static burden baseline and the single STDP transition model across the surveillance period (Fig. 2C). At the 72-hour prediction horizon, the calibrated STDP ensemble again outperformed the static burden baseline, with AUROC 0.838 [0.822-0.854] versus 0.800 [0.783-0.817], AUPRC 0.591 [0.553-0.627] versus 0.512 [0.476-0.553], and Brier score 0.150 [0.144-0.156] versus 0.170 [0.164-0.176] (Table 2; Supplementary Fig. S1). Thus, STDP-enhanced modeling improved both near-term and longer-horizon AKI prediction in a high-density ICU setting.

Feature attribution supported clinical interpretability of the STDP ensemble. High-gain contextual features included renal disease indicators, creatinine rise from baseline, central venous pressure measurement patterns, urine output rate, and early electrolyte measurements (Supplementary Fig. S1a). The strongest temporal transition signals reflected clinically plausible pathways linking cardiovascular disease, renal disease, urine-output abnormalities, fluid/electrolyte disorders, hypotension, lactate elevation, and hematologic abnormalities to subsequent AKI risk (Supplementary Fig. S1b,c). Transition-signal evolution analyses showed that several renal and cardiovascular transition patterns strengthened between the 24-to-48-hour and 24-to-72-hour horizons, consistent with accumulation of renal deterioration signals over time (Supplementary Fig. S1b). In the highest-risk decile, patients had greater creatinine rise, higher frequency of CVP-related measurements, more total laboratory measurements, and higher temperature measurement density, suggesting that the model captured both physiologic deterioration and increased clinical monitoring among patients at elevated AKI risk (Supplementary Fig. S1d).

### 3.4. PDAC early recurrence prediction

We next applied the STDP-inspired framework to early postoperative recurrence prediction in PDAC, a sparse longitudinal oncology task with irregular preoperative EHR observation. The primary clinical decision horizon was defined as Day –30 before surgery or index date, while Day 0 was retained as a sustained-performance horizon. In the rolling prediction cohort, 164 of 713 patients developed recurrence within 6 months. At Day –30, the calibrated STDP ensemble showed modestly higher discrimination than the static burden baseline, with AUROC 0.611 [95% CI, 0.560-0.659] versus 0.587 [0.534-0.636], AUPRC 0.323 [0.267-0.399] versus 0.318 [0.258-0.384], and Brier score 0.172 [0.157-0.189] versus 0.174 [0.158-0.190] (Fig. 3, Table 3). At Day 0, the calibrated STDP ensemble similarly showed higher performance than static burden, with AUROC 0.681 [0.636-0.728] versus 0.657 [0.610-0.705], AUPRC 0.363 [0.303-0.436] versus 0.331 [0.277-0.400], and Brier score 0.165 [0.149-0.181] versus 0.170 [0.154-0.187] (Table 3). These findings suggest that STDP-derived temporal transition features provided incremental information beyond cumulative preoperative burden, although the magnitude of improvement was modest.

**Figure 3.**
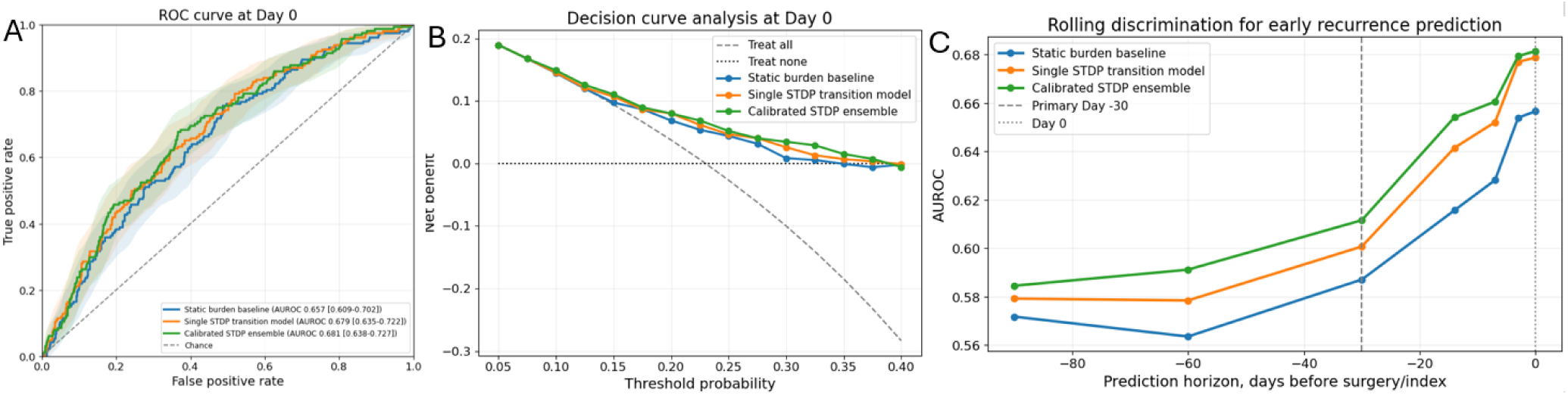
STDP-enhanced rolling prediction of early PDAC recurrence. A, ROC curves for postoperative recurrence prediction at Day 0. B, Decision-curve analysis at Day 0 comparing net benefit across threshold probabilities. C, Rolling AUROC across preoperative prediction horizons, with Day –30 marked as the primary decision horizon and Day 0 as the final preoperative snapshot. The calibrated STDP ensemble showed higher discrimination than the static burden baseline and single STDP transition model across rolling horizons.

**Table 3.**
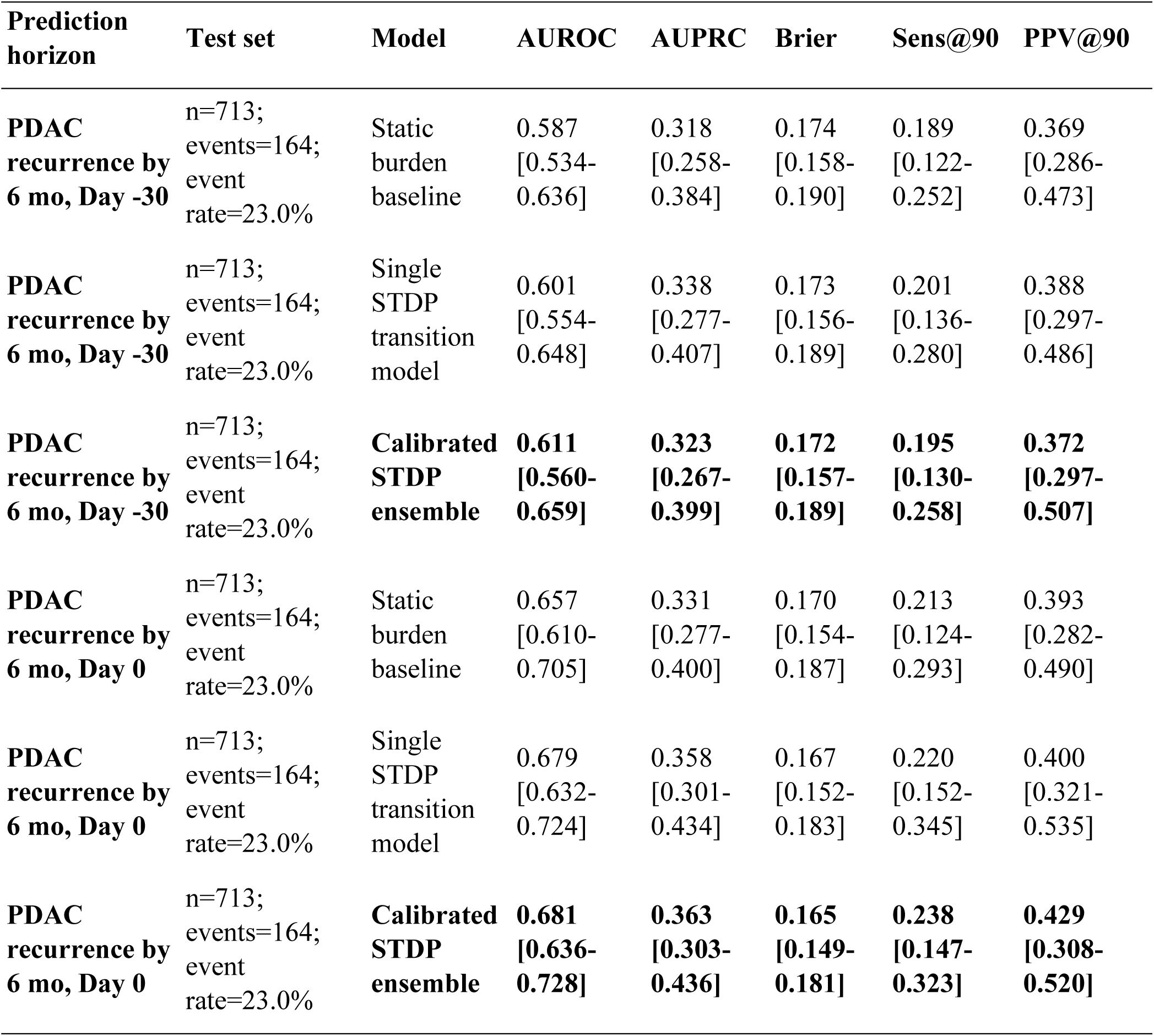
PDAC recurrence prediction performance at Day –30 and Day 0 horizons. Held-out rolling prediction performance for early postoperative PDAC recurrence within 6 months.

| Prediction horizon | Test set | Model | AUROC | AUPRC | Brier | Sens@90 | PPV@90 |
| --- | --- | --- | --- | --- | --- | --- | --- |
| **PDAC recurrence by 6 mo, Day -30** | n=713;<br>events=164;<br>event rate=23.0% | Static burden baseline | 0.587<br>[0.534-0.636] | 0.318<br>[0.258-0.384] | 0.174<br>[0.158-0.190] | 0.189<br>[0.122-0.252] | 0.369<br>[0.286-0.473] |
| **PDAC recurrence by 6 mo, Day -30** | n=713;<br>events=164;<br>event rate=23.0% | Single STDP transition model | 0.601<br>[0.554-0.648] | 0.338<br>[0.277-0.407] | 0.173<br>[0.156-0.189] | 0.201<br>[0.136-0.280] | 0.388<br>[0.297-0.486] |
| **PDAC recurrence by 6 mo, Day -30** | n=713;<br>events=164;<br>event rate=23.0% | **Calibrated STDP ensemble** | **0.611**<br>**[0.560-0.659]** | **0.323**<br>**[0.267-0.399]** | **0.172**<br>**[0.157-0.189]** | **0.195**<br>**[0.130-0.258]** | **0.372**<br>**[0.297-0.507]** |
| **PDAC recurrence by 6 mo, Day 0** | n=713;<br>events=164;<br>event rate=23.0% | Static burden baseline | 0.657<br>[0.610-0.705] | 0.331<br>[0.277-0.400] | 0.170<br>[0.154-0.187] | 0.213<br>[0.124-0.293] | 0.393<br>[0.282-0.490] |
| **PDAC recurrence by 6 mo, Day 0** | n=713;<br>events=164;<br>event rate=23.0% | Single STDP transition model | 0.679<br>[0.632-0.724] | 0.358<br>[0.301-0.434] | 0.167<br>[0.152-0.183] | 0.220<br>[0.152-0.345] | 0.400<br>[0.321-0.535] |
| **PDAC recurrence by 6 mo, Day 0** | n=713;<br>events=164;<br>event rate=23.0% | **Calibrated STDP ensemble** | **0.681**<br>**[0.636-0.728]** | **0.363**<br>**[0.303-0.436]** | **0.165**<br>**[0.149-0.181]** | **0.238**<br>**[0.147-0.323]** | **0.429**<br>**[0.308-0.520]** |

The magnitude of improvement in PDAC was smaller than in the AKI benchmark though significant relative to this cohort (Supplementary Fig. S2), consistent with the lower temporal density and longer disease timescale of oncology surveillance. Nevertheless, STDP-enhanced models showed consistent gains over static burden at Day –30 and Day 0 and retained modest decision-curve advantage across clinically relevant threshold ranges. Calibration analyses showed broadly similar predicted and observed recurrence risk across risk bins, with the calibrated ensemble reducing some instability seen in the single-transition and static models at rolling horizons (Supplementary Fig. S3). Feature attribution supported clinical interpretability: high-ranking static features included jaundice, obstructive gastrointestinal and hepatobiliary conditions, chronic pancreatitis, anemia, and liver disease, while transition features captured evolving hepatobiliary, nutritional, inflammatory, glycemic, renal, and symptom-related trajectories across preoperative horizons (Supplementary Fig. S4).

### 2.4. Robustness to heterogeneous EHR observation length and visit occurrences

Because STDP-derived features depend on observed temporal event pairs, we evaluated whether performance was driven by heterogeneous EHR record length or observation intensity. In representative PDAC robustness analyses, the calibrated STDP ensemble remained above the static burden baseline across increasing minimum prior-record requirements and within prior-record-length strata. At Day 0 prediction, observation intensity alone showed lower discrimination than clinical models, while STDP-enhanced models retained higher AUROC and AUPRC than observation-intensity-only models after adjustment (Fig. 4). These findings suggest that the STDP signal was not explained solely by longer EHR history, greater healthcare utilization, or denser documentation. Observation intensity alone had near-chance discrimination at Day –30, and STDP-based models retained higher AUROC and AUPRC than observation-intensity-only models, suggesting that the transition signal was not explained solely by encounter frequency or denser documentation (Supplementary Fig. S5). Because the AKI task occurred over a short ICU time scale, we evaluated observation heterogeneity using first-24-hour laboratory measurement intensity rather than long-term record length. The calibrated STDP ensemble showed consistently higher rolling AUPRC across ICU snapshots and remained competitive across low, medium, and high laboratory-intensity strata for both AUPRC and AUROC (Supplementary Fig. S6). Together, these analyses indicate that STDP-enhanced prediction was robust to heterogeneous observation patterns across both sparse longitudinal oncology data and dense ICU data, while also confirming that data density influences absolute model performance. We also explored whether aggregated STDP transition patterns could be summarized as a transition-velocity proxy, reflecting the relative concentration of temporally proximal transition signals across physiologic axes. This exploratory analysis highlighted liver/cholestatic, hematologic, glycemic, inflammatory, renal, and pancreatobiliary symptom axes as the strongest contributors, consistent with clinically plausible preoperative deterioration patterns (Supplementary Fig. S7). Together, these findings support the feasibility of STDP-enhanced rolling recurrence surveillance in a sparse within-disease prognostic task.

**Figure 4.**
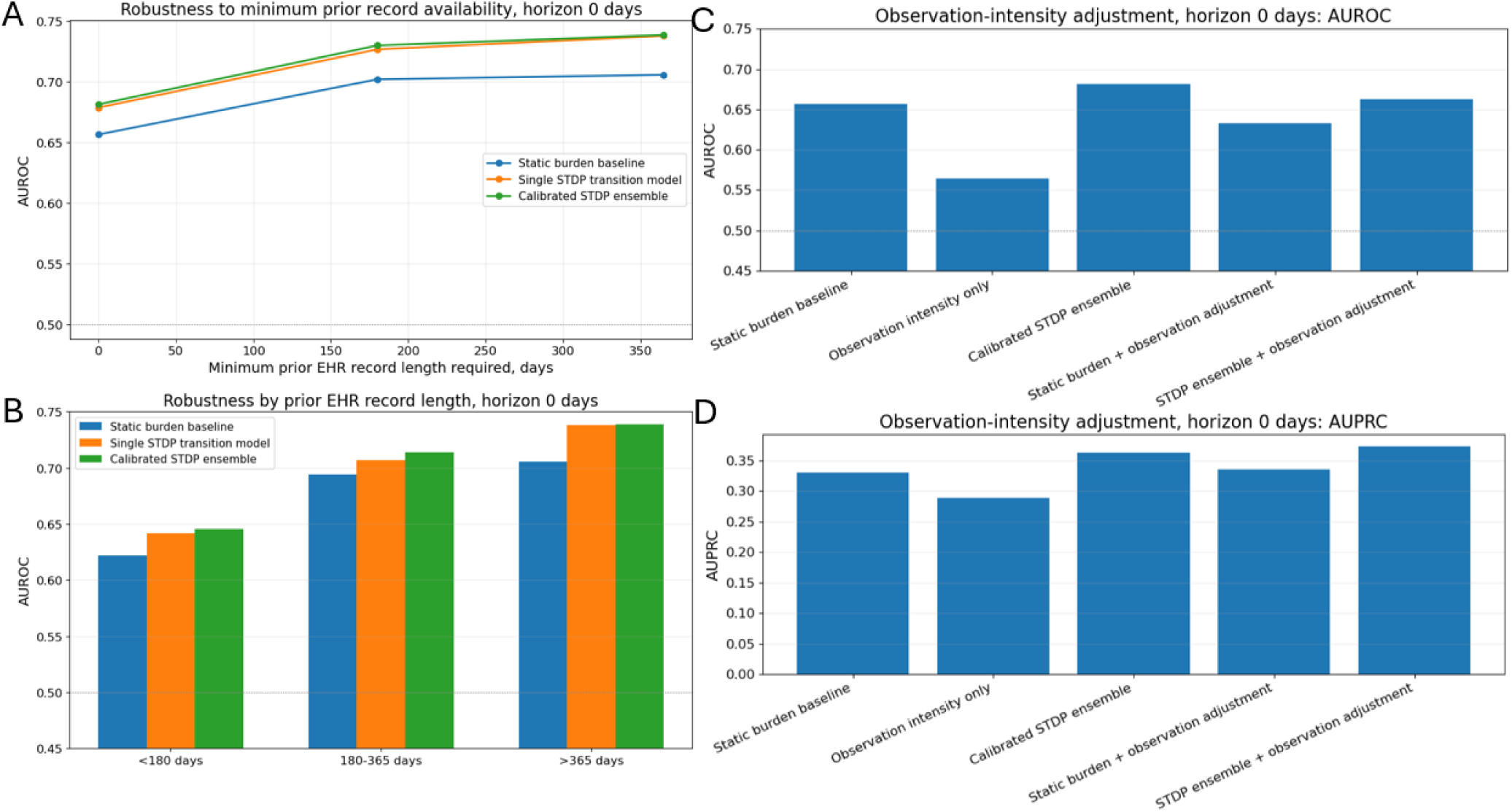
Robustness of PDAC recurrence prediction to EHR observation heterogeneity. A, AUROC after requiring increasing minimum prior EHR record availability before the prediction horizon. B, AUROC stratified by prior EHR record length. C,D, Observation-intensity adjustment at Day 0 using AUROC and AUPRC. Observation intensity alone showed lower discrimination than clinical models, while the calibrated STDP ensemble retained higher performance than static burden after adjustment.

## 4. Discussion

In this study, we developed and evaluated a simplified STDP-inspired temporal transition framework for adaptive risk prediction from structured EHR data. Rather than representing patient history only as cumulative burden or relying on latent sequential embeddings, the proposed approach converts irregular clinical histories into sparse, directional event-pair features. This representation preserves event identity, event order, and approximate temporal proximity while remaining compatible with standard supervised models and feature-level interpretation. The framework was evaluated across two clinically distinct settings: incident AKI prediction in MIMIC-IV, representing a dense ICU environment with frequent laboratory and physiologic measurements, and early postoperative PDAC recurrence prediction, representing a sparse longitudinal oncology surveillance task. Across these settings, STDP-enhanced models improved performance over static burden baselines, with larger gains in the dense AKI benchmark and more modest but consistent gains in the PDAC recurrence task. These findings support the central premise that temporal ordering can contribute predictive information beyond cumulative clinical burden, while also showing that the magnitude of benefit depends strongly on event density, outcome timescale, and the clinical setting.

The strongest performance gains were observed in AKI prediction, where the calibrated STDP ensemble improved AUROC, AUPRC, calibration, and high-specificity sensitivity at both 48-hour and 72-hour prediction horizons. This is consistent with the nature of AKI as an acute physiologic process that often unfolds over hours to days through temporally linked changes in renal function, urine output, hemodynamics, electrolytes, inflammatory markers, and clinical monitoring patterns. Prior EHR prediction studies have shown that temporal deep-learning models can improve clinical forecasting by learning sequential structure from longitudinal data, including recurrent and transformer-based representations.^10,11,14^ However, such models often require large datasets and encode temporal patterns in latent states that are difficult to inspect. In contrast, the STDP-inspired representation used here explicitly encodes directional transitions, allowing signals such as hemodynamic or cardiovascular context preceding renal deterioration to be examined directly. The AKI results therefore suggest that STDP-inspired feature construction may be particularly useful for short-horizon clinical deterioration tasks in which events are frequent, temporally proximal, and mechanistically related.

In PDAC recurrence prediction, the observed gains were smaller but clinically informative. The calibrated STDP ensemble modestly improved Day –30 preoperative recurrence prediction and showed similar incremental improvement at Day 0. This is a more difficult task than cancer-versus-control detection because all patients had the same underlying malignancy, and the model was asked to distinguish patients who developed recurrence within 6 months from those who remained recurrence-free. The smaller gain is consistent with sparse observation, heterogeneous surveillance patterns, and the longer biological timescale of recurrence. Nevertheless, the transition features highlighted clinically plausible hepatobiliary, inflammatory, glycemic, hematologic, renal, nutritional, and pancreatobiliary symptom axes. This suggests that the model captured temporally organized clinical deterioration patterns rather than only total code or laboratory burden. Importantly, these results should not be interpreted as sufficient for standalone surgical decision-making. Instead, the Day –30 horizon should be viewed as a clinically actionable risk-stratification setting in which a model could support tumor-board discussion, additional imaging, biomarker review, or intensified surveillance. The high-specificity evaluation was included for this reason: false-positive high-risk classification in PDAC could lead to unnecessary testing or altered treatment planning, so identifying a smaller subset of higher-risk patients while maintaining a low false-positive rate is clinically relevant.

This study has several limitations. First, EHR timestamps reflect both disease biology and clinical behavior, including documentation patterns, visit frequency, ordering practices, and surveillance intensity. Although robustness analyses suggested that STDP performance was not explained solely by record length or observation intensity, residual confounding by healthcare utilization remains possible. Second, the STDP equations used here are a computational analogy adapted from pair-based spike-timing-dependent plasticity, not a mechanistic model of neurobiological plasticity. Lag and decay parameters were empirical hyperparameters rather than fixed biological constants. Although shorter windows improved performance in selected models, further tuning and external validation are needed to determine whether these parameters identify clinically meaningful transition windows or primarily optimize prediction within a given dataset. Third, the PDAC cohort was derived from a single institution and requires external validation before clinical generalization. Fourth, the current implementation used structured EHR features and simplified transition families; future work should evaluate integration with imaging, pathology, medications, free-text reports, and prospective workflows. Finally, although STDP-inspired features provide greater interpretability than many latent sequential models, they should still be interpreted as temporally informed associations rather than causal pathways. Future applications may be especially valuable in even shorter-timescale settings, such as emergency department deterioration, sepsis, stroke, or vital-sign-driven prediction, where event timing may carry stronger signal and tuned lag windows could help define clinically useful monitoring intervals.

## 5. Conclusions

We developed an STDP-inspired temporal transition framework that represents asynchronous EHR histories as sparse, directional, and interpretable event-pair features. Across two prediction settings, the approach improved AKI prediction in a dense ICU benchmark and provided modest but interpretable gains for early PDAC recurrence prediction in a sparse longitudinal oncology cohort. The framework offers a practical alternative to purely static burden models and more opaque sequential deep-learning approaches by preserving temporal ordering while retaining feature-level interpretability. Further external validation and prospective evaluation are needed to determine whether STDP-enhanced rolling risk prediction can support clinical decision-making across acute and chronic care settings.

## Data and Code availability

MIMIC-IV data are available through PhysioNet to credentialed users who complete the required training and agree to the applicable data-use agreement. The CUMC institutional EHR data used in this study cannot be publicly shared because they contain protected health information and are subject to HIPAA regulations, institutional policies, and data-use restrictions. To support reproducibility while maintaining compliance, the analytic code has been cleaned to remove any institution-specific paths, credentials, identifiers, or protected health information and is available at: https://github.com/abundanceli

## Acknowledgements

ChatGPT (OpenAI) was used for editorial assistance, including language refinement and polishing of selected manuscript text. All scientific content, interpretation, and final wording were reviewed and approved by the authors. The authors thank the Columbia University Herbert Irving Comprehensive Cancer Center for its support.

## Funding

This work was supported in part by the Columbia University Herbert Irving Comprehensive Cancer Center through the National Cancer Institute of the National Institutes of Health under award number P30 CA013696.

## Author contributions

Liyuan Gong (Conceptualization, Data curation, Formal analysis, Investigation, Methodology, Project administration, Supervision, Validation, Visualization, Writing-original draft, Writing-review & editing), Nitish Aswani (Data curation, Validation, Writing-review & editing), Jeong Yun, Yang (Validation, Writing-review & editing), Peter Shahinnian (Validation, Writing-review & editing), Despina Kontos (Validation, Writing-review & editing) Gulam Manji (Validation, Writing-review & editing), Stella Kang (Validation, Writing-review & editing), and Chin Hur (Funding acquisition, Investigation, Supervision, Writing-review & editing)

## Competing interests

The authors declare no competing interest.

## Supplementary materials

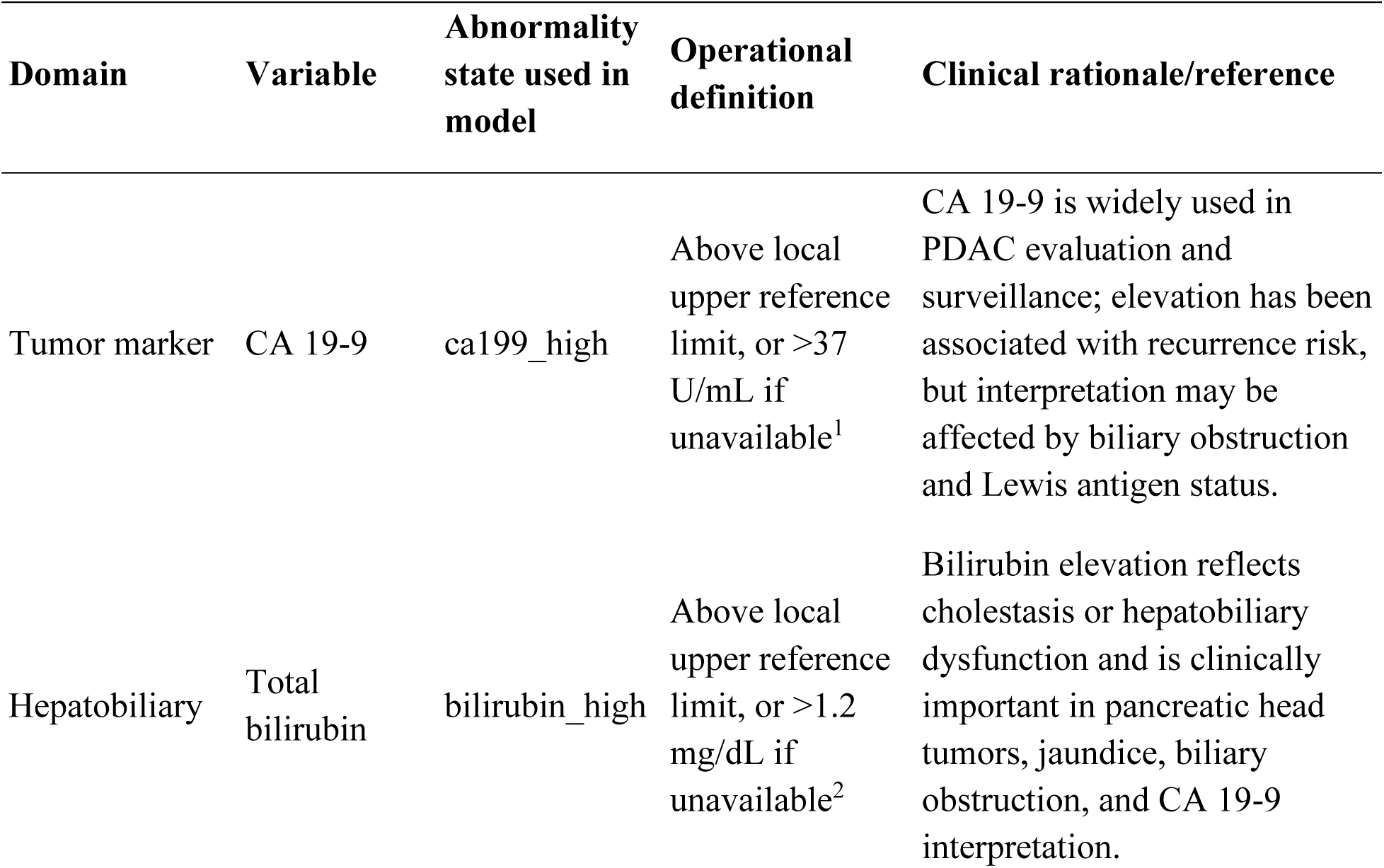
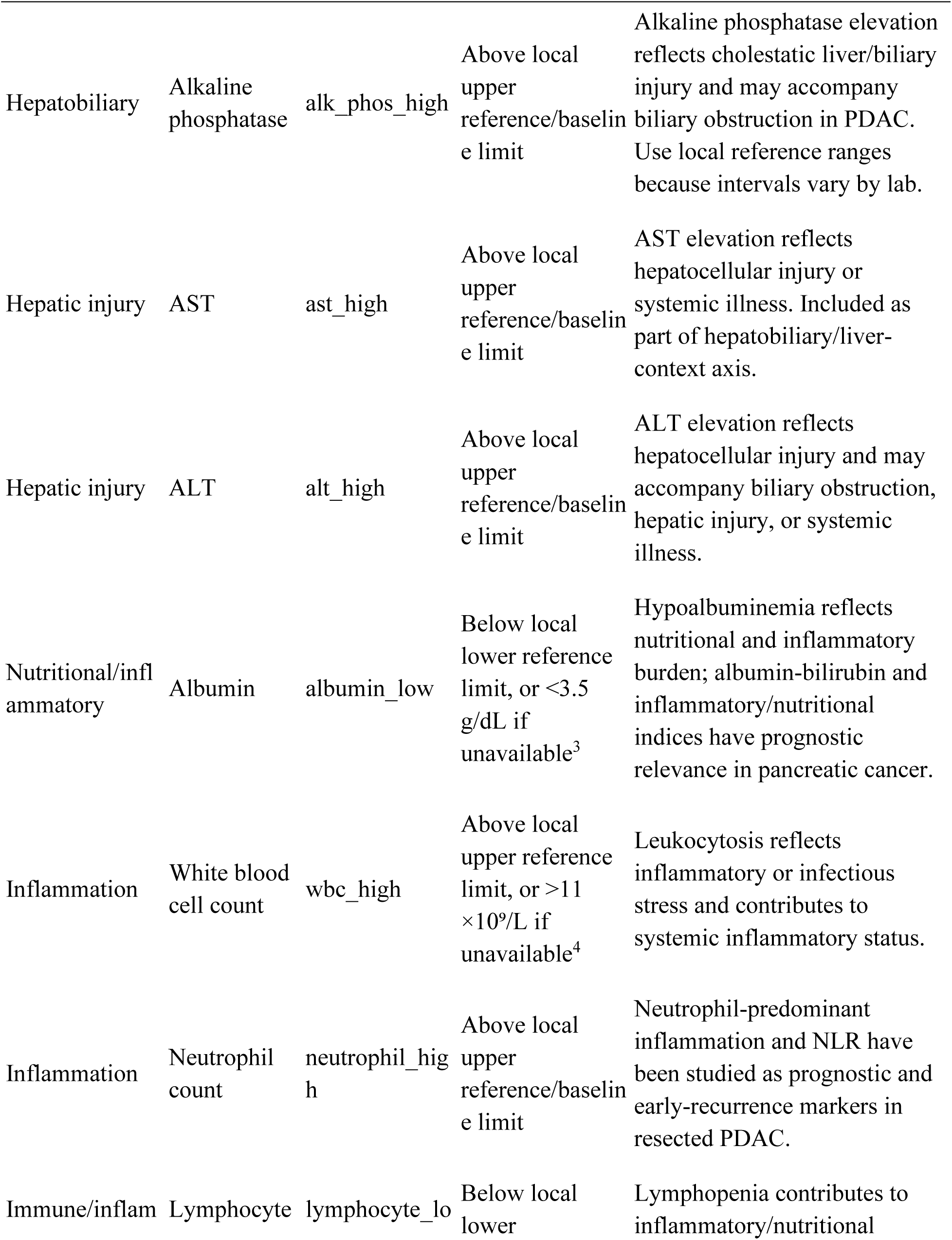
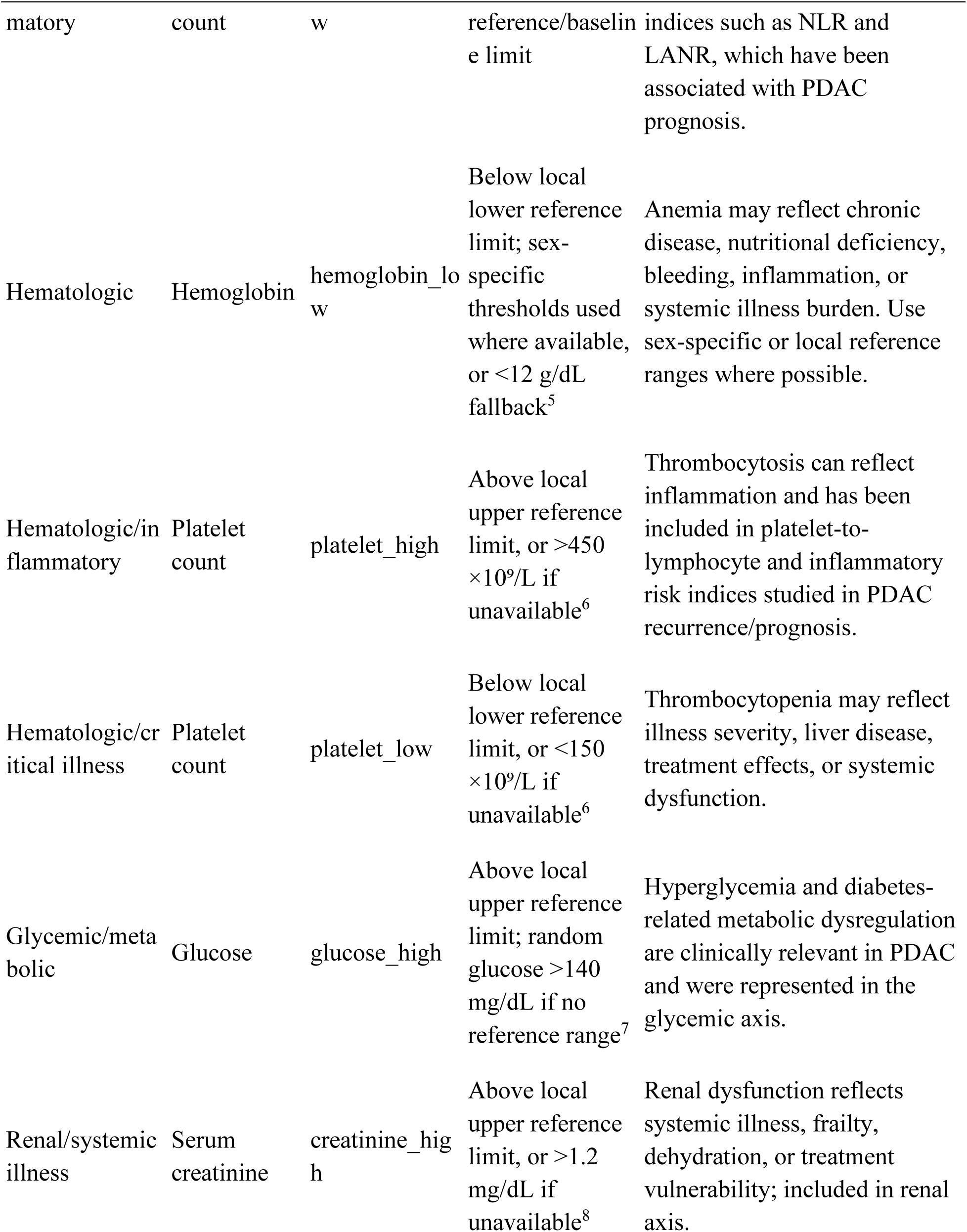
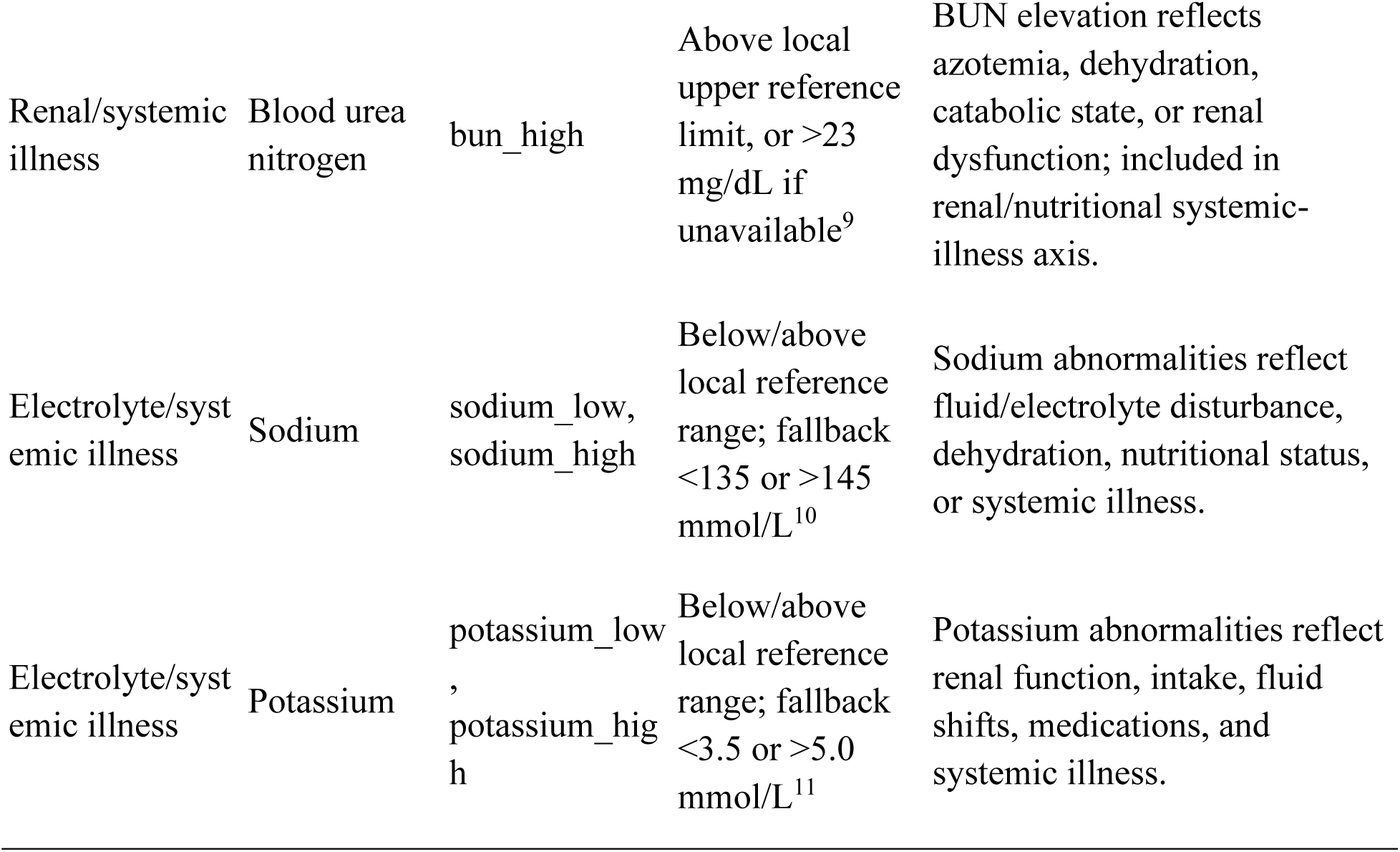
Table S1. **Supplementary Table S1 footnote**. Laboratory abnormality states were defined using local laboratory reference ranges when available. When local ranges were unavailable, widely used adult fallback thresholds were applied. These thresholds were used for feature engineering and physiologic-axis grouping, not for clinical diagnosis. CA 19-9 was interpreted cautiously because elevation can occur with biliary obstruction, cholangitis, inflammation, and Lewis antigen non-secretor status, and recurrence assessment in PDAC should not rely on CA 19-9 alone.

**Figure S1.**
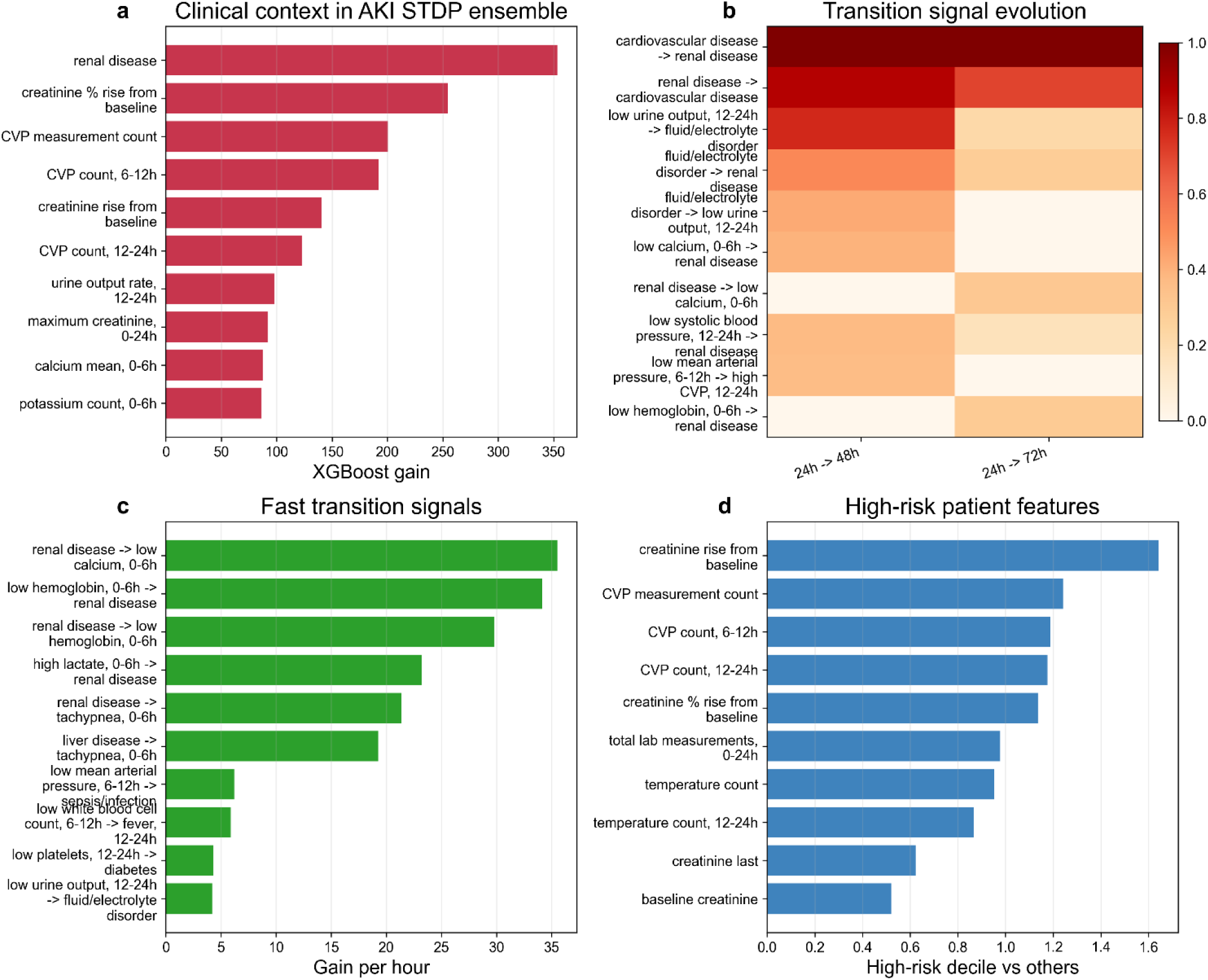
Feature attribution and temporal transition signals in the AKI STDP ensemble. a, Top clinical context features ranked by XGBoost gain. b, Evolution of selected STDP transition signals between the 24-to-48-hour and 24-to-72-hour prediction horizons. c, Fast transition signals ranked by gain per hour. d, Features enriched in the highest-risk decile compared with remaining patients. The highlighted features and transitions reflect clinically plausible renal, hemodynamic, electrolyte, inflammatory, and hematologic patterns associated with incident AKI risk.

**Figure S2.**
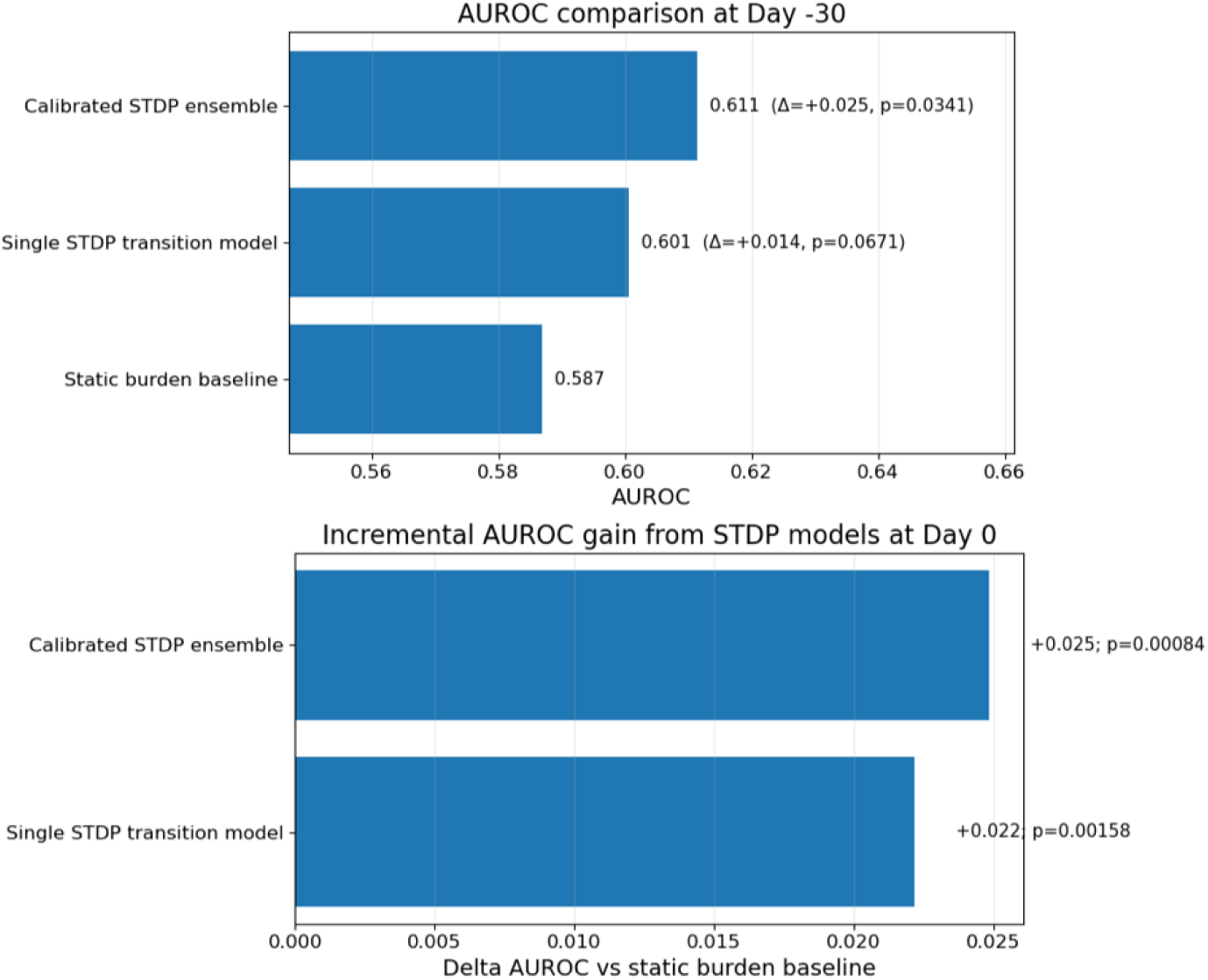
Paired DeLong comparison of PDAC recurrence models. AUROC comparisons between static burden, single STDP transition, and calibrated STDP ensemble models for PDAC recurrence prediction. At Day –30, the calibrated STDP ensemble showed a modest AUROC gain over static burden, while the single STDP transition model showed a smaller, borderline difference. At Day 0, both STDP-based models showed statistically significant AUROC gains over the static burden baseline by paired DeLong testing.

**Figure S3.**
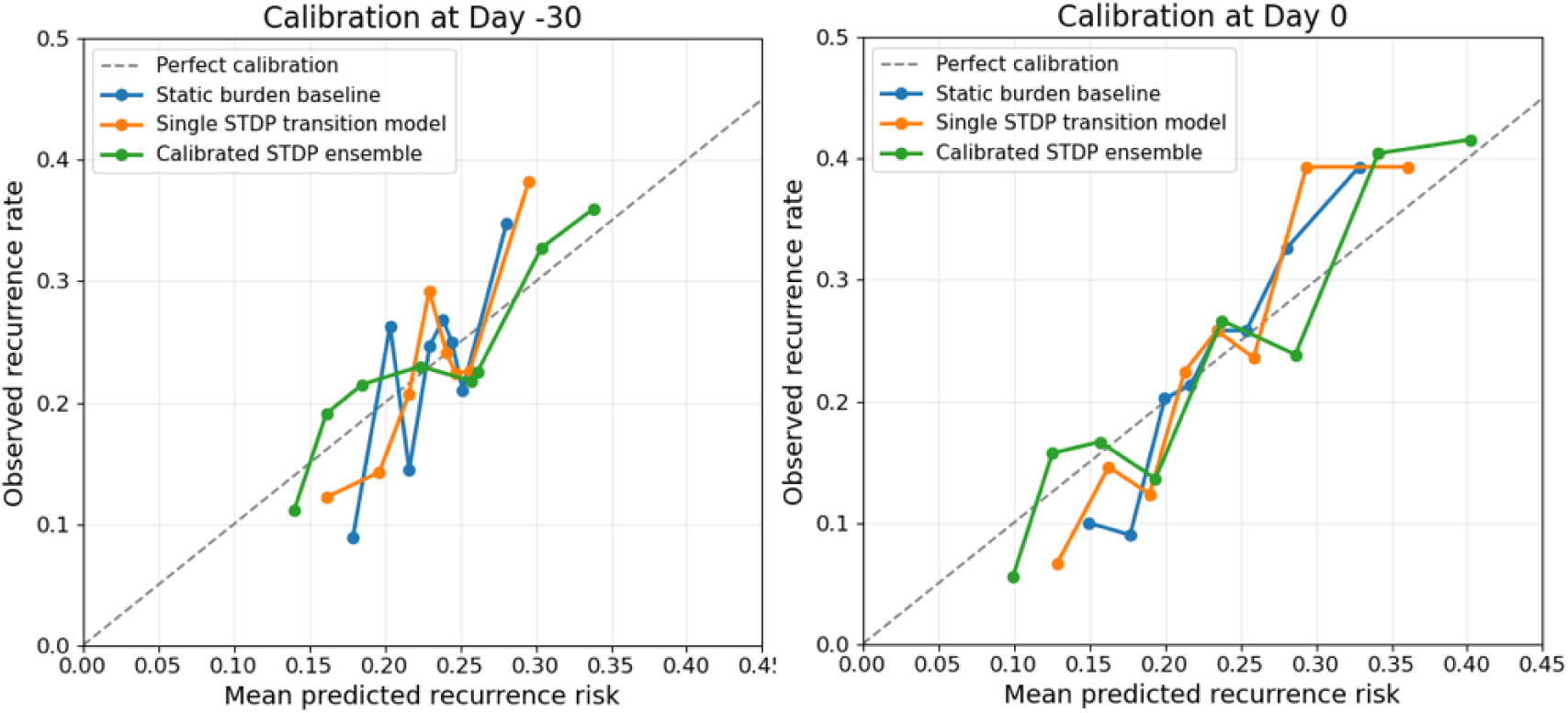
Calibration of PDAC recurrence prediction at Day –30 and Day 0. Calibration curves compare predicted and observed 6-month recurrence risk for the static burden baseline, single STDP transition model, and calibrated STDP ensemble. The dashed diagonal line indicates perfect calibration. Across both horizons, predicted risk generally increased with observed recurrence rate, with some variability across risk bins reflecting the modest cohort size and sparse recurrence events.

**Figure S4.**
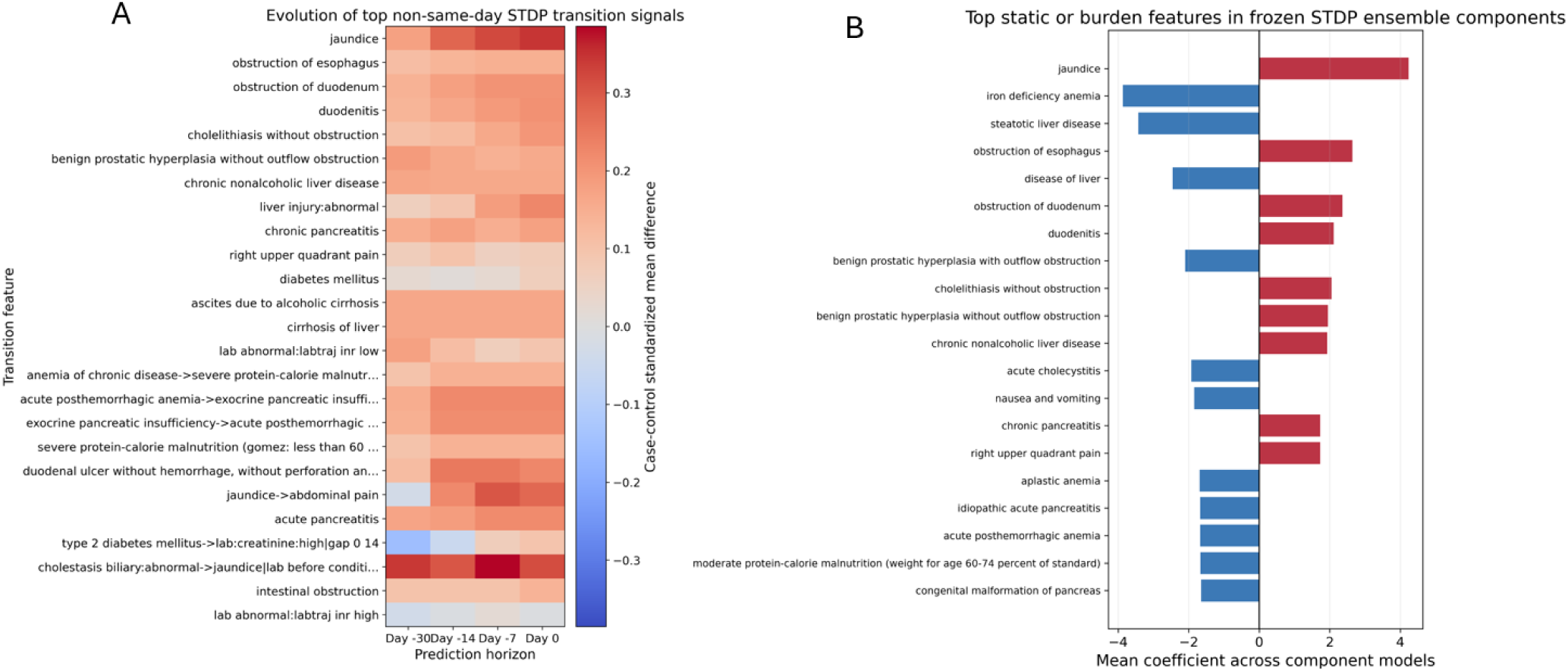
PDAC transition-signal evolution and static feature attribution. A, Evolution of top non-same-day STDP transition signals across preoperative prediction horizons. Color indicates the standardized case-control mean difference for each transition feature. B, Top static or burden features in frozen STDP ensemble components, ranked by mean coefficient across component models. Positive coefficients indicate features associated with higher predicted recurrence risk, whereas negative coefficients indicate features associated with lower predicted recurrence risk.

**Figure S5.**
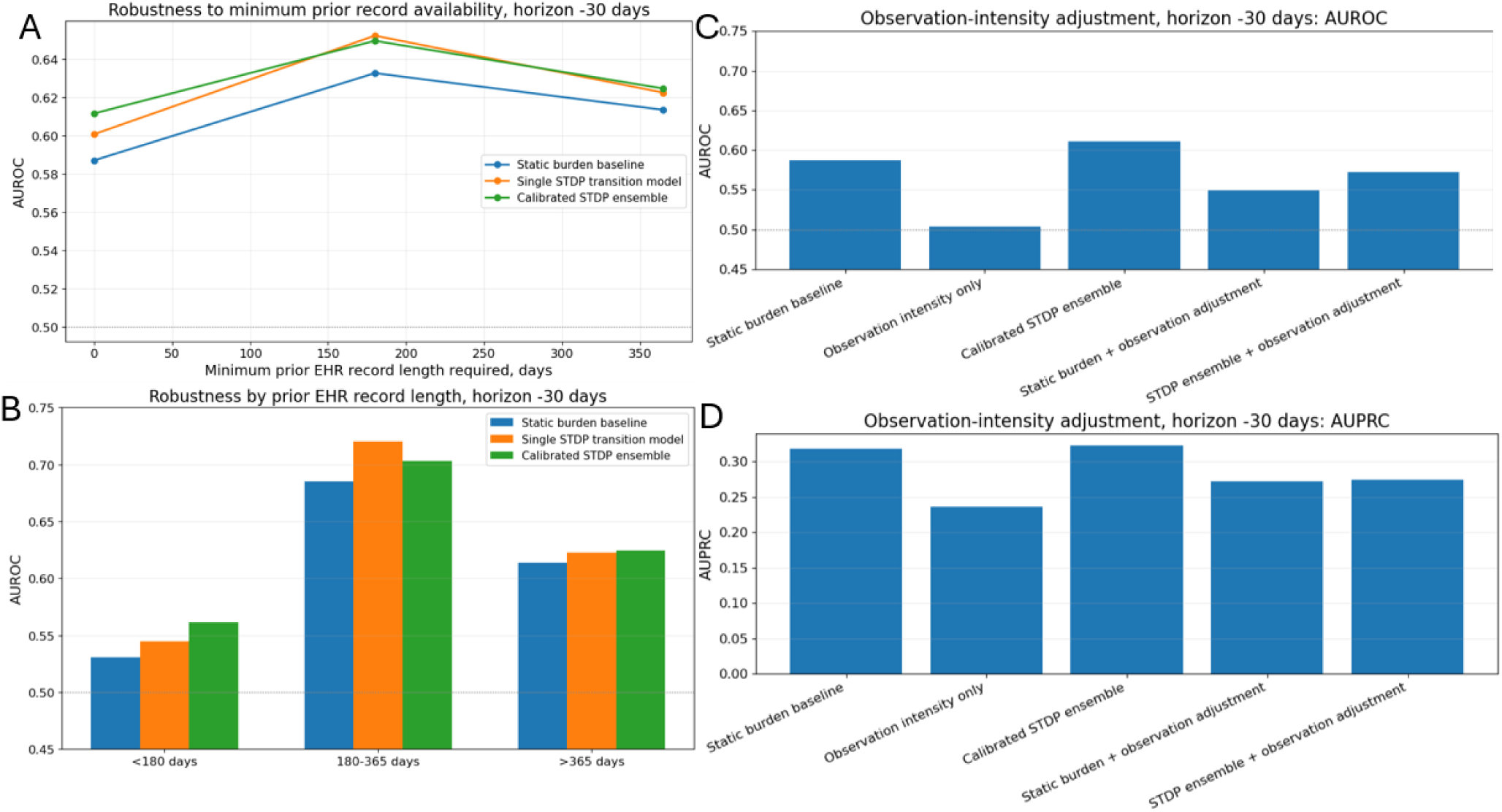
Robustness of PDAC Day –30 recurrence prediction to EHR record length and observation intensity. A, AUROC after requiring increasing minimum prior EHR record availability before the prediction horizon. B, AUROC stratified by prior EHR record length. C,D, Observation-intensity adjustment at Day –30 using AUROC and AUPRC. Observation intensity alone showed limited discrimination, while STDP-enhanced models retained higher performance than observation-intensity-only models.

**Figure S6.**
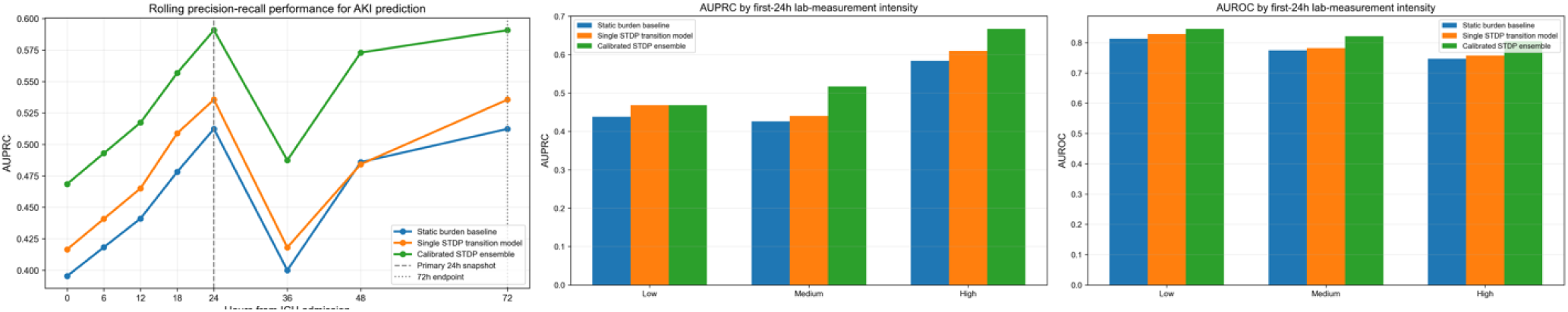
Robustness of AKI prediction to laboratory measurement intensity. Rolling AUPRC and stratified AUPRC/AUROC are shown across first-24-hour laboratory measurement-intensity strata. The calibrated STDP ensemble maintained higher precision-recall performance across ICU snapshots and showed comparable or higher discrimination across low, medium, and high laboratory-intensity groups, supporting robustness of the AKI transition signal to differences in measurement density.

**Figure S7.**
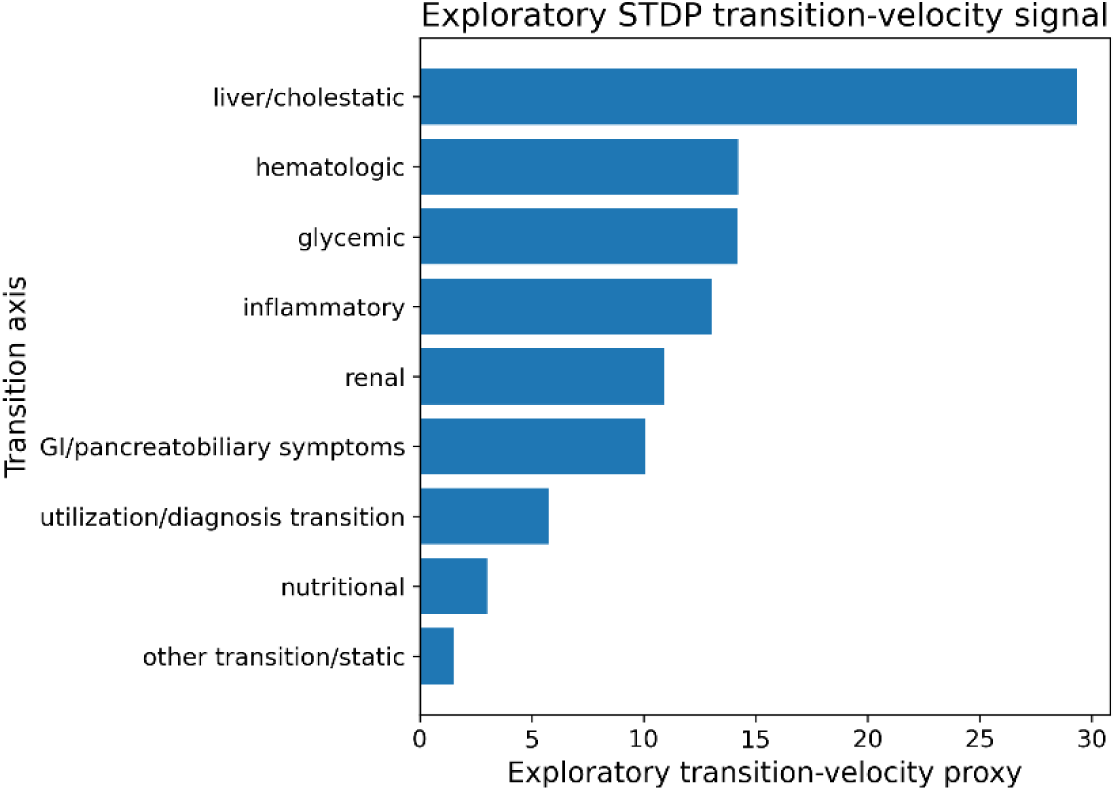
Exploratory STDP transition-velocity proxy in PDAC recurrence prediction. Coefficient-weighted STDP transition signals were aggregated by grouped physiologic axes to estimate an exploratory transition-velocity proxy. Higher values indicate axes with a greater concentration of temporally proximal transition signals contributing to recurrence-risk prediction. Liver/cholestatic, hematologic, glycemic, inflammatory, renal, and gastrointestinal/pancreatobiliary symptom axes showed the strongest signals.

## Notes

### Competing Interest Statement

The authors have declared no competing interest.

### Author Declarations

Ethics committee/IRB of Columbia University Irving Medical Center gave ethical approval for this work. The approved IRB protocol number was AAAS1319

