## supplementary materials for "STDP-inspired temporal transition modeling for adaptive clinical risk prediction from electronic health records"

Table S1

| Domain | Variable | Abnormality state used in model | Operational definition | Clinical rationale/reference |
|---|---|---|---|---|
| Tumor marker | CA 19-9 | ca199_high | Above local upper reference limit, or >37 U/mL if unavailable[1] | CA 19-9 is widely used in PDAC evaluation and surveillance; elevation has been associated with recurrence risk, but interpretation may be affected by biliary obstruction and Lewis antigen status. |
| Hepatobiliary | Total bilirubin | bilirubin_high | Above local upper reference limit, or >1.2 mg/dL if unavailable[2] | Bilirubin elevation reflects cholestasis or hepatobiliary dysfunction and is clinically important in pancreatic head tumors, jaundice, biliary obstruction, and CA 19-9 interpretation. |
| Hepatobiliary | Alkaline phosphatase | alk_phos_high | Above local upper reference/baseline limit | Alkaline phosphatase elevation reflects cholestatic liver/biliary injury and may accompany biliary obstruction in PDAC. Use local reference ranges because intervals vary by lab. |
| Hepatic injury | AST | ast_high | Above local upper reference/baseline limit | AST elevation reflects hepatocellular injury or systemic illness. Included as part of hepatobiliary/liver-context axis. |
| Hepatic injury | ALT | alt_high | Above local upper reference/baseline limit | ALT elevation reflects hepatocellular injury and may accompany biliary obstruction, hepatic injury, or systemic illness. |

| Domain | Variable | Abnormality state used in model | Operational definition | Clinical rationale/reference |
|---|---|---|---|---|
| Nutritional/inflammatory | Albumin | albumin_low | Below local lower reference limit, or <3.5 g/dL if unavailable[3] | Hypoalbuminemia reflects nutritional and inflammatory burden; albumin-bilirubin and inflammatory/nutritional indices have prognostic relevance in pancreatic cancer. |
| Inflammation | White blood cell count | wbc_high | Above local upper reference limit, or >11 $\times 10^9$/L if unavailable[4] | Leukocytosis reflects inflammatory or infectious stress and contributes to systemic inflammatory status. |
| Inflammation | Neutrophil count | neutrophil_high | Above local upper reference/baseline limit | Neutrophil-predominant inflammation and NLR have been studied as prognostic and early-recurrence markers in resected PDAC. |
| Immune/inflammatory | Lymphocyte count | lymphocyte_low | Below local lower reference/baseline limit | Lymphopenia contributes to inflammatory/nutritional indices such as NLR and LANR, which have been associated with PDAC prognosis. |
| Hematologic | Hemoglobin | hemoglobin_low | Below local lower reference limit; sex-specific thresholds used where available, or <12 g/dL fallback[5] | Anemia may reflect chronic disease, nutritional deficiency, bleeding, inflammation, or systemic illness burden. Use sex-specific or local reference ranges where possible. |
| Hematologic/inflammatory | Platelet count | platelet_high | Above local upper reference limit, or >450 | Thrombocytosis can reflect inflammation and has been included in platelet-to-lymphocyte and inflammatory |

| Domain | Variable | Abnormality state used in model | Operational definition | Clinical rationale/reference |
|---|---|---|---|---|
| | | | $\times 10^9$/L if unavailable[6] | risk indices studied in PDAC recurrence/prognosis. |
| Hematologic/critical illness | Platelet count | platelet_low | Below local lower reference limit, or <150 $\times 10^9$/L if unavailable[6] | Thrombocytopenia may reflect illness severity, liver disease, treatment effects, or systemic dysfunction. |
| Glycemic/metabolic | Glucose | glucose_high | Above local upper reference limit; random glucose >140 mg/dL if no reference range[7] | Hyperglycemia and diabetes-related metabolic dysregulation are clinically relevant in PDAC and were represented in the glycemic axis. |
| Renal/systemic illness | Serum creatinine | creatinine_high | Above local upper reference limit, or >1.2 mg/dL if unavailable[8] | Renal dysfunction reflects systemic illness, frailty, dehydration, or treatment vulnerability; included in renal axis. |
| Renal/systemic illness | Blood urea nitrogen | bun_high | Above local upper reference limit, or >23 mg/dL if unavailable[9] | BUN elevation reflects azotemia, dehydration, catabolic state, or renal dysfunction; included in renal/nutritional systemic-illness axis. |
| Electrolyte/systemic illness | Sodium | sodium_low, sodium_high | Below/above local reference range; fallback <135 or >145 mmol/L[10] | Sodium abnormalities reflect fluid/electrolyte disturbance, dehydration, nutritional status, or systemic illness. |
| Electrolyte/systemic illness | Potassium | potassium_low, potassium_high | Below/above local reference range; fallback | Potassium abnormalities reflect renal function, intake, fluid shifts, medications, and systemic illness. |

| Domain | Variable | Abnormality state used in model | Operational definition | Clinical rationale/reference |
|---|---|---|---|---|
| | | | <3.5 or >5.0 mmol/L[11] | |

**Supplementary Table S1 footnote.** Laboratory abnormality states were defined using local laboratory reference ranges when available. When local ranges were unavailable, widely used adult fallback thresholds were applied. These thresholds were used for feature engineering and physiologic-axis grouping, not for clinical diagnosis. CA 19-9 was interpreted cautiously because elevation can occur with biliary obstruction, cholangitis, inflammation, and Lewis antigen non-secretor status, and recurrence assessment in PDAC should not rely on CA 19-9 alone.

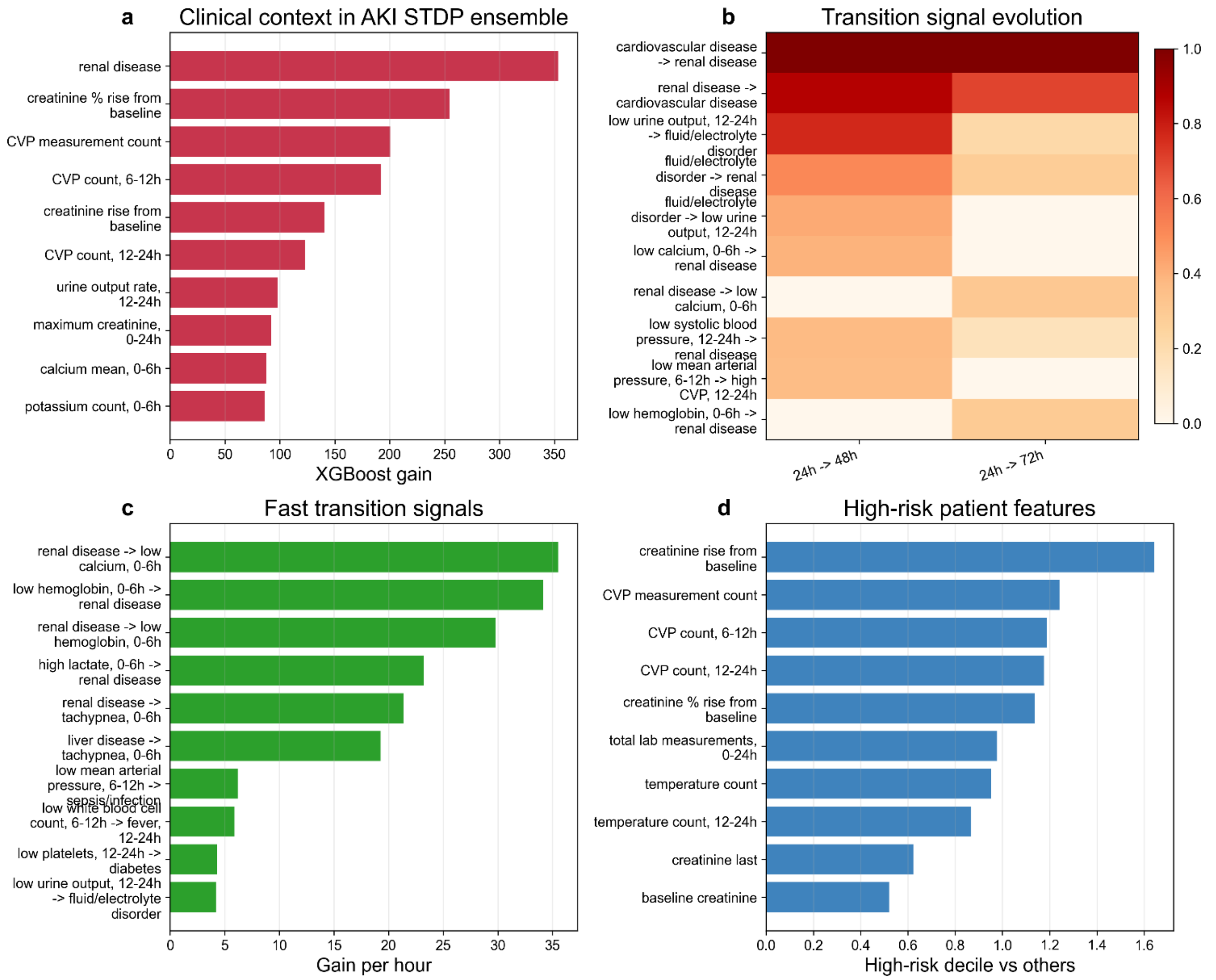


**Figure S1 Feature attribution and temporal transition signals in the AKI STDP ensemble.** a, Top clinical context features ranked by XGBoost gain. b, Evolution of selected STDP transition signals between the 24-to-48-hour and 24-to-72-hour prediction horizons. c, Fast transition signals ranked by gain per hour. d, Features enriched in the highest-risk decile compared with remaining patients. The highlighted features and transitions reflect clinically plausible renal, hemodynamic, electrolyte, inflammatory, and hematologic patterns associated with incident AKI risk.

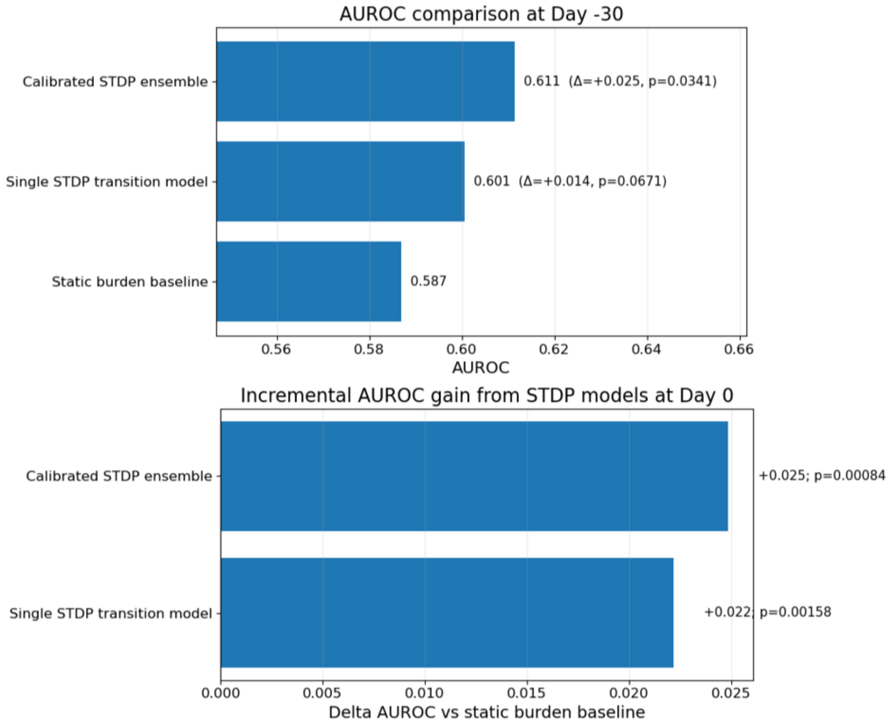


**Figure S2 Paired DeLong comparison of PDAC recurrence models.** AUROC comparisons between static burden, single STDP transition, and calibrated STDP ensemble models for PDAC recurrence prediction. At Day -30, the calibrated STDP ensemble showed a modest AUROC gain over static burden, while the single STDP transition model showed a smaller, borderline difference. At Day 0, both STDP-based models showed statistically significant AUROC gains over the static burden baseline by paired DeLong testing.

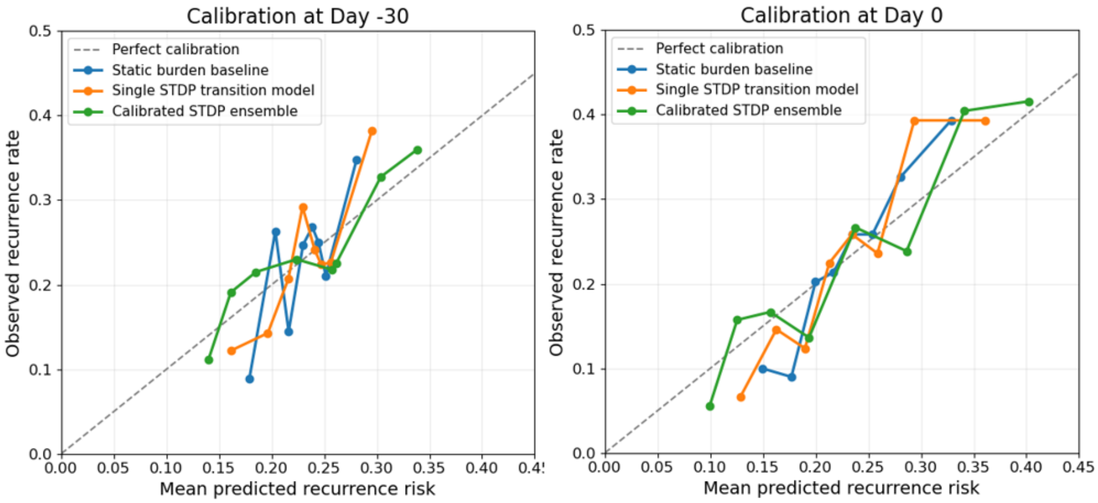


**Figure S3 Calibration of PDAC recurrence prediction at Day -30 and Day 0.** Calibration curves compare predicted and observed 6-month recurrence risk for the static burden baseline, single STDP transition model, and calibrated STDP ensemble. The dashed diagonal line indicates perfect calibration. Across both horizons, predicted risk generally increased with observed recurrence rate, with some variability across risk bins reflecting the modest cohort size and sparse recurrence events.

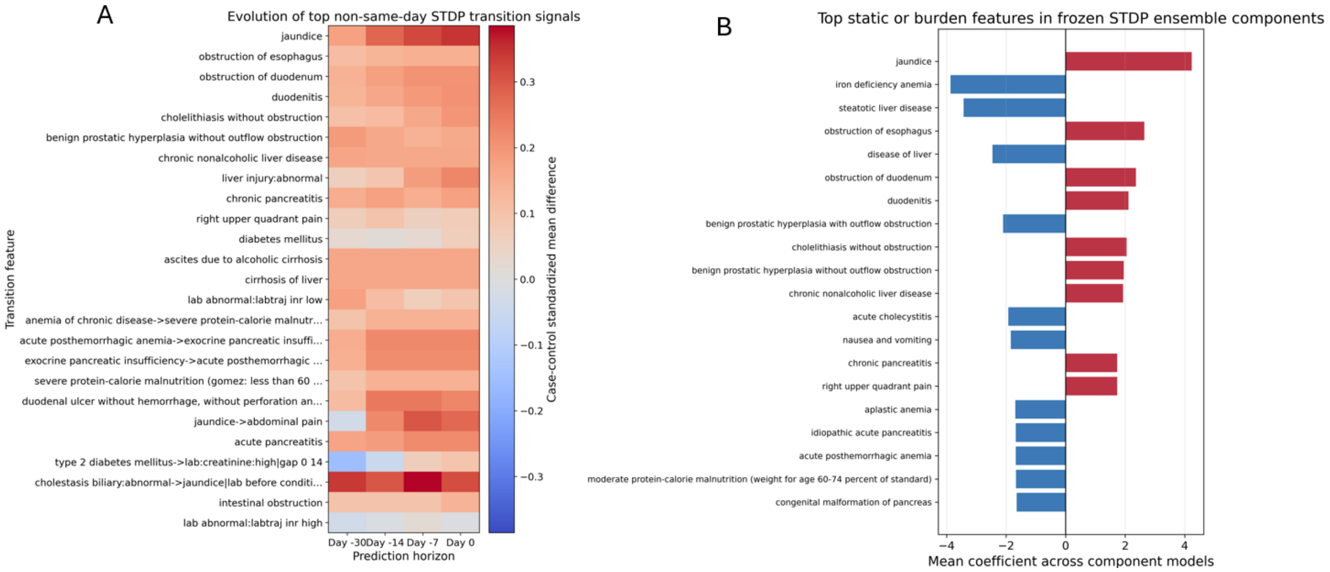


**Figure S4 PDAC transition-signal evolution and static feature attribution.** A, Evolution of top non-same-day STDP transition signals across preoperative prediction horizons. Color indicates the standardized case-control mean difference for each transition feature. B, Top static or burden features in frozen STDP ensemble components, ranked by mean coefficient across component models. Positive coefficients indicate features associated with higher predicted recurrence risk, whereas negative coefficients indicate features associated with lower predicted recurrence risk.

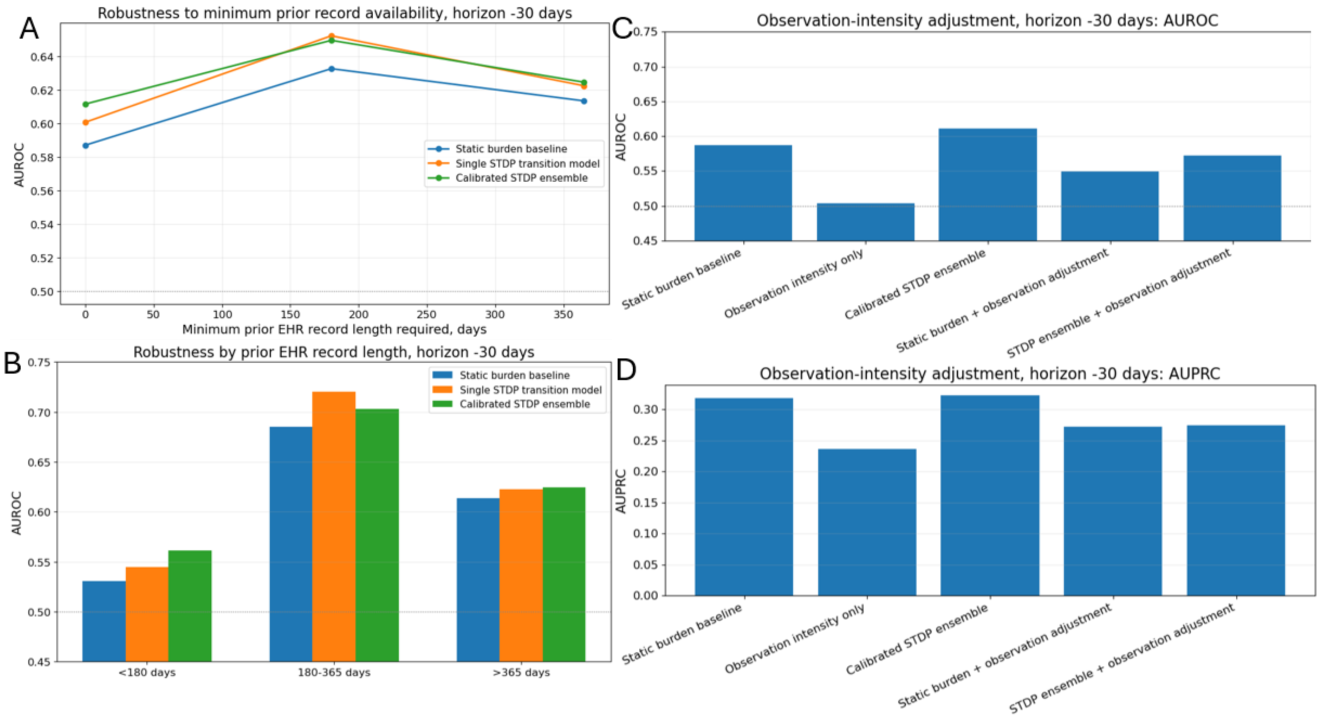


**Figure S5 Robustness of PDAC Day -30 recurrence prediction to EHR record length and observation intensity.** A, AUROC after requiring increasing minimum prior EHR record availability before the prediction horizon. B, AUROC stratified by prior EHR record length. C,D, Observation-intensity adjustment at Day -30 using AUROC and AUPRC. Observation intensity alone showed limited discrimination, while STDP-enhanced models retained higher performance than observation-intensity-only models.

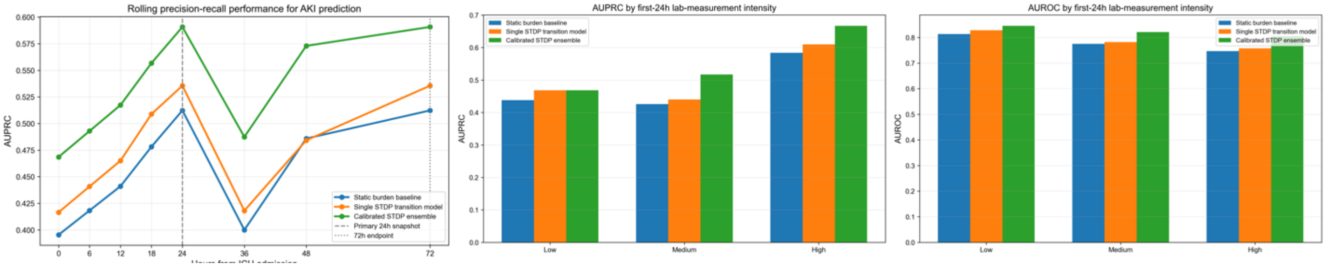


**Figure S6 Robustness of AKI prediction to laboratory measurement intensity.** Rolling AUPRC and stratified AUPRC/AUROC are shown across first-24-hour laboratory measurement-intensity strata. The calibrated STDP ensemble maintained higher precision-recall performance across ICU snapshots and showed comparable or higher discrimination across low, medium, and high laboratory-intensity groups, supporting robustness of the AKI transition signal to differences in measurement density.

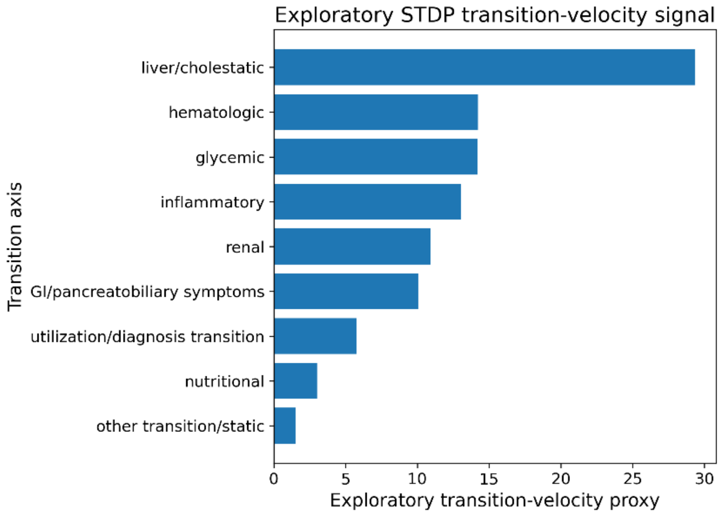


**Figure S7 Exploratory STDP transition-velocity proxy in PDAC recurrence prediction.** Coefficient-weighted STDP transition signals were aggregated by grouped physiologic axes to estimate an exploratory transition-velocity proxy. Higher values indicate axes with a greater concentration of temporally proximal transition signals contributing to recurrence-risk prediction. Liver/cholestatic, hematologic, glycemic, inflammatory, renal, and gastrointestinal/pancreatobiliary symptom axes showed the strongest signals.
